# Evaluating the role of amino acids in type 2 diabetes risk: a Mendelian randomization study

**DOI:** 10.1101/2023.08.27.23294702

**Authors:** Jacky Man Yuen Mo, Baoting He, Tommy Hon Ting Wong, Ying Liang, Shan Luo, Kenneth Lo, Jimmy Chun Yu Louie, Shiu Lun Au Yeung

## Abstract

**Background:** Previous observational and Mendelian randomization studies suggested different amino acids associated with type 2 diabetes (T2D). However, these studies may suffer from confounding or the use of invalid instruments, respectively.

**Methods:** We extracted strong (*p* < 5×10^-8^), independent (r^2^ < 0.001) genetic variants associated with nine amino acids (alanine, glutamine, glycine, histidine, phenylalanine, tyrosine, isoleucine, leucine, and valine) from summary statistics of UK Biobank (N ≤ 115,075), with exclusion of potentially pleiotropic variants. We then applied them to T2D summary statistics from DIAMANTE Consortium (without UK Biobank participants) (N = 455,313) and FinnGen study (N = 365,950), and glycemic traits (MAGIC consortium, N ≤ 209,605). Inverse variance weighed (IVW) method was the main analysis, with multiple sensitivity analyses to assess robustness of findings.

**Results:** Alanine was associated with higher T2D risk, correcting for multiple testing (Odds Ratio (OR) 1.50 per SD; 95% CI 1.16 to 1.95). At nominal significance, isoleucine was associated with higher T2D risk (OR 1.13; 95% CI 1.00 to 1.27) and tyrosine was associated with lower T2D risk (OR 0.89; 95% CI 0.80 to 0.99). Alanine was also associated with lower insulin, higher glycated hemoglobin and glucose whereas isoleucine and leucine were associated with lower insulin. These associations were consistent in most sensitivity analyses.

**Conclusion:** Alaine likely contributed to higher T2D risk whilst the associations for isoleucine and tyrosine requires further verification. Whether these findings explain health effects of sources of amino acids, such as diet, should be further explored.

## Introduction

Type 2 diabetes (T2D) is projected to affect over 570 million adults worldwide by 2025, with an increasing prevalence and associated burden on healthcare systems across the globe.^1^ As such, identifying modifiable etiologic factors related to T2D would aid prevention and early diagnosis of T2D. Recent research have increasingly suggested certain amino acids, in particularly branched chain amino acids, are potential risk factors of T2D. Earlier prospective cohort studies suggest alanine, tyrosine and isoleucine may increase T2D whilst glycine and serine inversely associated with T2D risk.^2–4^ Although the exact mechanistic pathways were not fully elucidated, impairment of insulin secretion and insulin resistance may be one of these pathways.^5 6^ However, whether these findings were causal were unclear given the possibility of confounding by socioeconomic position and lifestyle factors in these observational studies.

Mendelian randomization (MR), a design that is less vulnerable to confounding than conventional observational studies,^7^ have been increasingly used to ascertain the causal role of metabolomic signatures associated with T2D. Previous MR studies have shown branch-chained amino acids (BCAAs), including leucine, isoleucine and valine, may increase T2D risk whereas alanine and tyrosine, an aromatic amino acid, may decrease T2D.^8–10^ However, these MR studies may be flawed due to the use of limited number of instruments derived from small genome wide association studies (GWAS) of amino acids,^11 12^ and the use of a single instrument (e.g. variants in protein phosphatase Mg^2+^/Mn^2+^ dependent 1K [*PPM1K*] locus) to estimate the causal relationship between multiple amino acids and T2D and hence constitute horizontal pleiotropy.^9 13^ In order to better evaluate the role of these amino acids in T2D risk taking into account of potentially invalid instruments in early MR studies, we conducted one of the largest MR studies to date by leveraging large GWAS summary statistics of nine amino acids covered in the Nuclear Magnetic Resonance (NMR) spectrometry platform in UK Biobank,^14^ with exclusion of pleiotropic instruments based on NMR measured metabolites,^15 16^ and respective GWAS of T2D and glycemic traits in Europeans.

## Methods

A valid MR study should fulfill three instrumental variable assumptions.^17^ First, relevance – genetic instruments are strongly associated with the exposure of interest. Second, independence – the instruments are free from confounding between the exposure and outcome. Third, exclusion restriction – the effect of instruments on the outcome must be exerted via the exposure with no alternative pathways.

### Genetic instruments of amino acids

The metabolic profiling of plasma samples from a randomly subset of participants (N = 118,461) from the UK Biobank was quantified using high-throughput NMR spectroscopy metabolomics (Nightingale Health Ltd., Helsinki, Finland, https://nightingalehealth.com/).^18^ We obtained strong (*p* value < 5×10^-8^), independent (r^2^ < 0.001) genetic predictors, single nucleotide polymorphism (SNPs), of nine circulating amino acids including BCAAs (isoleucine, leucine, and valine), aromatic amino acids (phenylalanine and tyrosine) and other amino acids (alanine, glutamine, glycine, and histidine) (**Supplementary Table 1)**, generated by Integrative Epidemiology Unit, University of Bristol (https://gwas.mrcieu.ac.uk/).^19^ Measurements of amino acids were inverse rank-normalized and the genetic associations of amino acids were obtained using a linear mixed model adjusted for age, sex, fasting status, and genotyping array. To reduce the impact of horizontal pleiotropy which biases MR estimates, we removed instruments that are associated with more than five NMR metabolites (including non-amino acid metabolites) in the same UK Biobank summary statistics, as per previous studies (**Supplementary Figure 1**).^15 16^

### Genetic association of type 2 diabetes

We obtained genetic association from the DIAbetes Meta-Analysis of Trans-Ethnic association studies (DIAMANTE) consortium, which is the largest genetic consortium of T2D at the time of analysis. To avoid possible bias arising from overlap of samples used in the amino acid GWAS and the T2D GWAS, we restricted the analyses without the UK Biobank samples, which included 55,005 cases and 400,308 controls of European.^20^ T2D was defined differently across studies, such as blood sugar level (glycated hemoglobin [HbA_1c_] ≥ 6.5%, fasting glucose [FG] ≥ 7.0 mmol/L or 2-hour glucose [2hGlu] ≥ 11.1 mmol/L), the use of anti-diabetic medication and physician diagnosis. The genetic associations were obtained using multivariable linear regression, adjusted for age, sex, and the top 10 genetic principal components (PC).

We also obtained genetic associations of T2D from FinnGen study (Round 9), a combination of Finnish Biobanks, which consists of 57,698 T2D cases and 308,252 controls participants.^21^ T2D was defined based on hospital record linkage and the genetic associations of T2D were obtained using multivariable linear regression adjusted for age, sex and the top 10 PCs.

### Genetic association of glycemic traits

We obtained genetic associations of FI (log-transformed), HbA_1c_ (%), FG (mmol/L) and 2hGlu (mmol/L) from the Meta-Analyses of Glucose and Insulin-related traits Consortium (MAGIC), restricted to participants of European descent (N ≤ 209,605).^22^ Participants with type 1 or type 2 diabetes, who reported taking anti-diabetic medications or had FG ≥ 7 mmol/L, 2hGlu ≥ 11.1 mmol/L, or HbA_1c_ ≥ 6.5% were excluded.^22^ The genetic associations of glycemic traits were obtained using multivariable linear regression adjusted for age, sex, body mass index (except for HbA_1c_), study-specific covariates and PCs. Although adjustment for covariable in GWAS may introduce collider bias, the original GWAS investigators did not find strong evidence of such bias.^22 23^

### Main analysis

We assessed the instrument strength using F statistic via approximation, where F statistic >10 indicated low evidence for weak instrument bias.^24^ We calculated the variance explained (R^2^) of each amino acid by summing up R^2^ of individual instrument, which was calculated using effect allele frequencies, effect estimates and corresponding standard errors, as per a previous study.^25 26^ We obtained the causal estimates of amino acids with T2D using inverse-variance weighted (IVW) method with multiplicative random effects, which assumed no overall horizontal pleiotropy.^27^ We assessed I^2^ for instrument heterogeneity where a high I^2^ indicated the possibility of invalid instruments. To increase the statistical power, we meta-analyzed the IVW estimates from DIAMANTE and FinnGen using random effect meta-analysis.

### Sensitivity analyses

To triangulate the evidence from MR methods which relied on different assumptions, we also conducted MR-Egger, which allows for genetic pleiotropy but requires the instrument strength independent of direct effect (InSIDE) assumption;^28^ weighted median method, which assumes at least half of weights were derived from valid instruments,^29^ and MR-Pleiotropy RESidual Sum and Outlier (PRESSO), which removed possible invalid instruments via the MR-PRESSO outlier test.^30^ We assessed overall horizontal pleiotropy using MR-Egger intercept test.^28^

To correct for multiple testing whilst taking into account the correlated nature of the amino acid, we calculated the principal components (PCs) needed to explain 99% of the variance of the nine amino acid exposures, using individual level data of our ongoing UK Biobank project (Application number: 14864), which gave 7 PCs. Statistical significance was therefore set as p < 0.0071 (0.05/7).

All analyses were performed using “TwoSampleMR”, “MendelianRandomization” and “MR--PRESSO” package on R (version 4.3.1).

### Ethics approval

This study only used publicly available summary data and hence no ethics approval is required. Details of ethical approval and participant consent for each of the studies that contributed to the GWAS can be found in the original publications.^19–22^

## Results

**Supplementary Table 2** shows the list of instruments for each amino acid. All instruments had an F statistic of >10, indicating low level of evidence for weak instruments bias. The F-statistics and R^2^ for each amino acid are as follow: alanine (30-250, 1.91%), glutamine (28-2209, 4.51%), glycine (26-377, 3.16%), histidine (35-157, 0.77%), phenylalanine (30-664, 1.13%), tyrosine (30-1027, 2.63%), isoleucine (33-267, 0.60%), leucine (32-488, 0.93%) and valine (30-673, 1.58%). Variants which were excluded due to associations with more than 5 metabolites were shown in **Supplementary Table 3**.

### Association of genetically predicted circulating amino acids with risk of type 2 diabetes

Figure 1 shows the association between genetically predicted amino acids and T2D risk. After combining the IVW estimates from DIAMENTE and FinnGen, per standard deviation (SD) increase in alanine (Odds ratio (OR) 1.50; 95% CI 1.16 to 1.95) was associated with higher T2D risk, correcting for multiple testing. At nominal significance, histidine (OR 1.32; 95% CI 1.04 to 1.66) and isoleucine (OR 1.13; 95% CI 1.00 to 1.27) were associated with higher T2D risk. Conversely, genetically predicted tyrosine (OR 0.89; 95% CI 0.80 to 0.99) was associated with lower T2D risk. These findings were generally consistent across both DIAMENTE and FinnGen (**Supplementary Table 4**).

**Figure 1:**
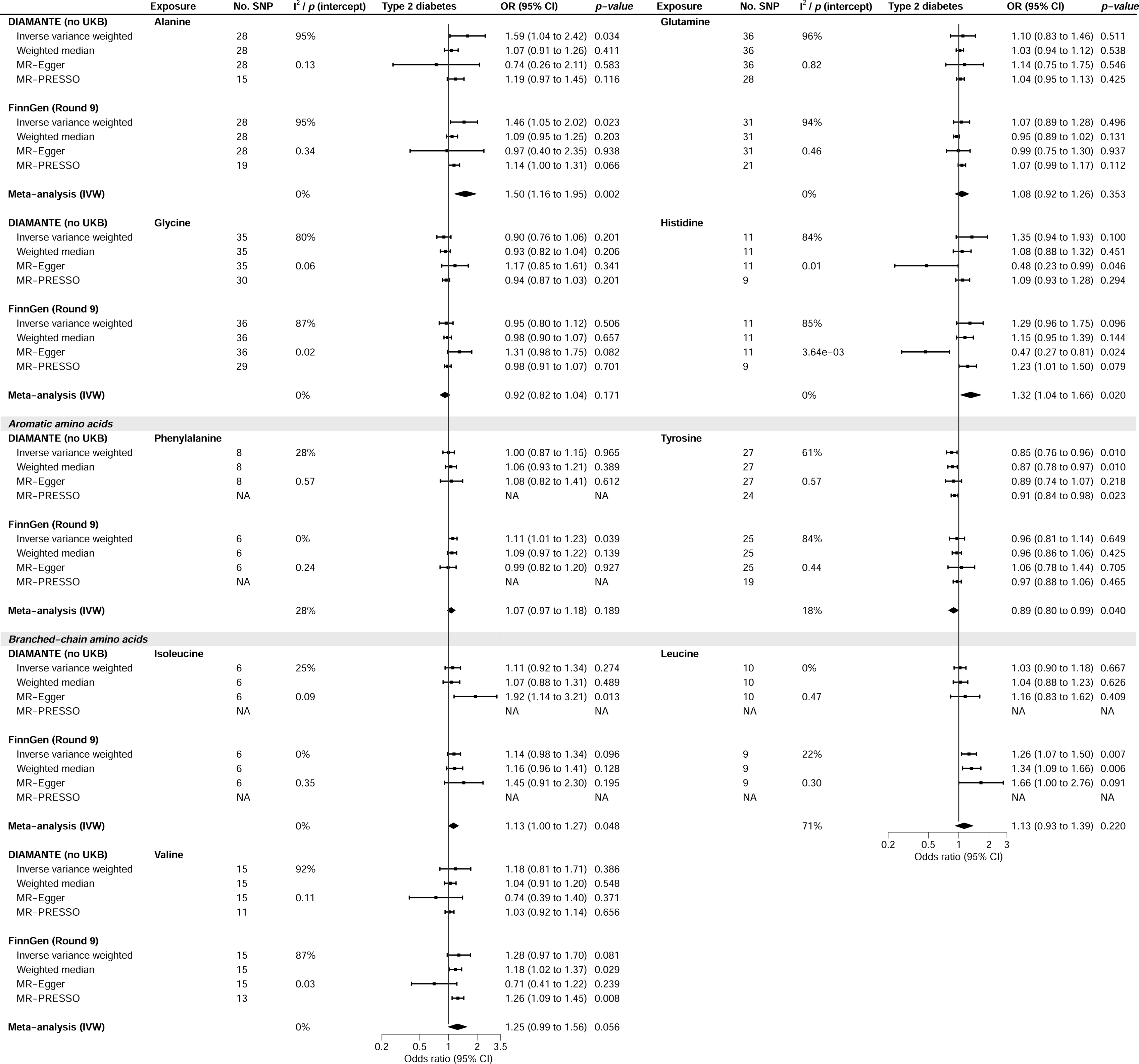
The association of genetically predicted amino acids (per SD) with type 2 diabetes (T2D) risk

**Supplementary Table 5** and **6** show the MR analyses using DIAMANTE and FinnGen, respectively. There were signs of heterogeneity based on *I*^2^ for most analyses. Notably, horizontal pleiotropy was evident for histidine, implying possible biases in IVW. Sensitivity analyses gave directionally similar estimates apart from MR-Egger regarding alanine- and histidine-associated analyses.

### Association of genetically predicted circulating amino acids with glycemic traits

Figures 2 and **3** show the association of genetically predicted amino acids with glycemic traits, including FI, FG, HbA_1c_ and 2hGlu. Using inverse-variance weighted, alanine (Beta [β]-0.06; 95% CI −0.09 to −0.03) and two BCAAs (isoleucine [β −0.07; 95% CI −0.11 to −0.03] and leucine [β −0.05; 95% CI −0.08 to −0.02]) were associated with lower FI, (Figure 2, left panel). Alanine was also associated with higher HbA_1c_ (β 0.06; 95% CI 0.02 to 0.11) (Figure 2, right panel) and FG (β 0.14; 95% CI 0.06 to 0.23) (Figure 3, left panel). There were signs of heterogeneity although evidence for horizontal pleiotropic effects was limited. Sensitivity analyses generally gave directionally similar estimates (**Supplementary Table 7-10**).

**Figure 2:**
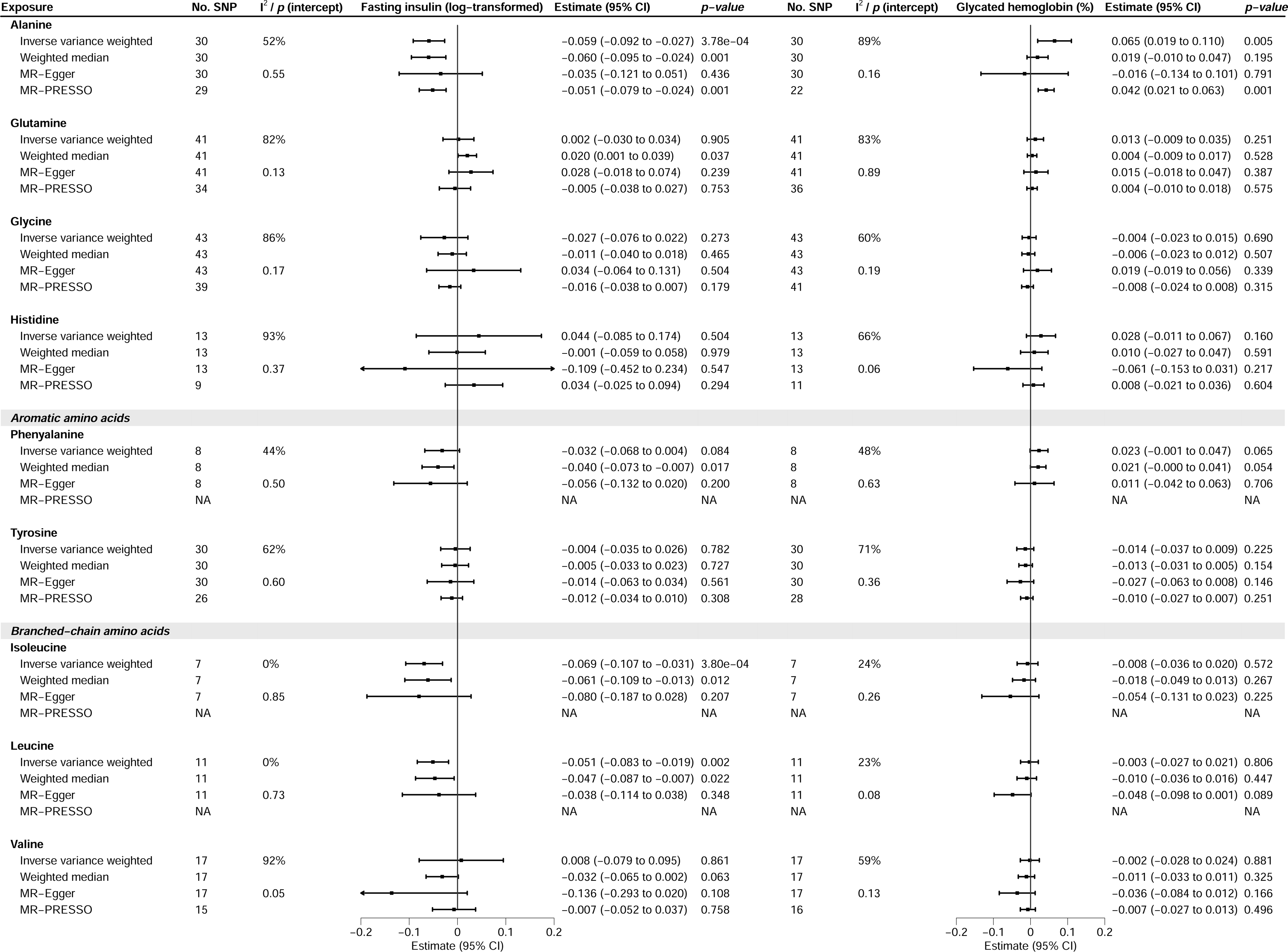
The association of genetically predicted amino acids (per SD) with fasting insulin and glycated hemoglobin

**Figure 3:**
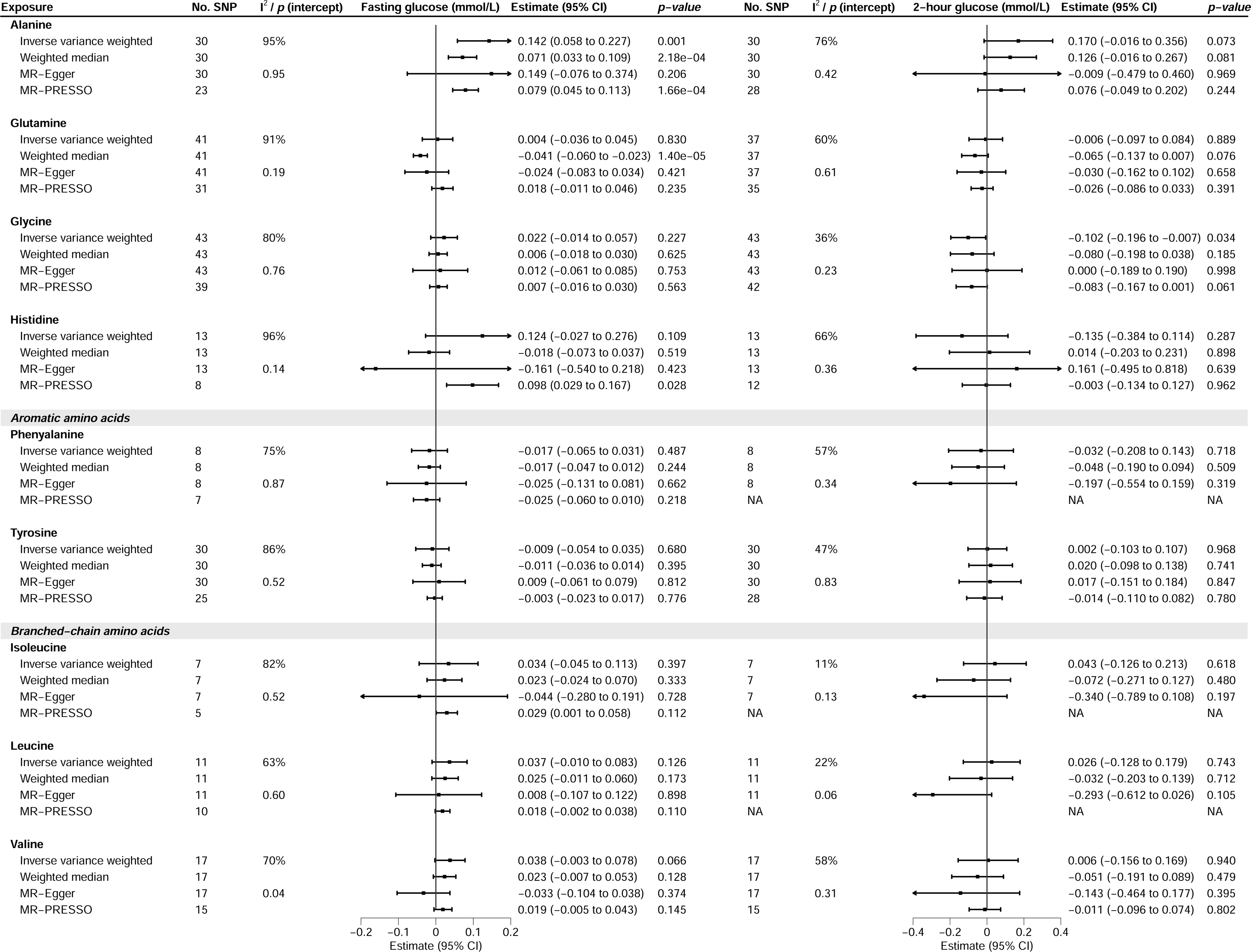
The association of nine genetically predicted amino acids (per SD) with fasting glucose and 2-hour glucose

## Discussion

To the best of our knowledge, this is one of the first MR studies which comprehensively assessed the role of nine amino acids in T2D risk and glycemic traits using respective large genetic consortia where earlier MR studies have only investigated BCAAs and tyrosine, and may suffer from methodological issues such as the use of smaller amino acid GWAS, the use of a single variant for investigation of multiple BCAAs.^8–10^ We showed, for the first-time using MR, that alanine likely increases T2D risk, and hence confirmed the positive findings in previous observational studies.^31–33^ Consistent with previous studies, our study showed that isoleucine (one of the BCAAs) likely increases T2D risk.^9 34^ However, the positive association between histidine and T2D risk was likely due to horizontal pleiotropy. In addition, we found tyrosine, an aromatic amino acid, to be inversely associated with T2D risk, which was consistent with a previous MR study but not observational studies.^8 35^

The positive association between alanine and glycemia concurred with the findings from earlier observational studies,^31–33^ and our study extended by showing an increased risk of T2D with higher alanine using a design less vulnerable to confounding. This is in contrary to an earlier study which showed an inverse association of alanine on T2D risk based on a single variant ^10^ which is vulnerable to horizontal pleiotropy. The potential harm of alanine in glucose homeostasis was also consistent with its association with increased fasting glucose and reduced insulin, consistent with a previous prospective cohort study.^2^ Although how alanine impacts T2D risk has not been thoroughly explored, alanine is involved in the glucose-alanine cycle where alanine is converted into pyruvate by alanine aminotransferase in the liver and then to glucose via the gluconeogenic pathway.^36 37^ Hence, it is postulated that elevated alanine may lead to hyperglycemia and reduce insulin secretion, subsequently induces insulin resistance,^38^ a hallmark feature of T2D. Alternatively, other studies suggested elevated alanine amongst people with non-alcoholic fatty liver disease,^32 39^ which in itself likely increases T2D risk.^40^ Additional studies would be necessary to clarify the role of alanine in these two related diseases. Our study also helped clarify that phenylalanine unlikely increases T2D risk, in contrary to a small prospective cohort study.^41^

The positive association of BCAAs with T2D risk was well studied in observational studies.^3 35 41 42^ With the inclusion of more genetic instruments, our study clarified that amongst the BCAAs, isoleucine may be more relevant to the development of T2D, consistent with findings from various analyses and data sources,^9^ although it is noted that the association did not pass correction for multiple testing and hence requires replications. Nevertheless, previous studies suggested that BCAAs could lead to persistent mammalian target of rapamycin complex 1 (mTORC1) activation, causing insulin resistance.^5^ On the other hand, others speculated that elevated BCAAs reflects impaired BCAA metabolism and hence the accumulation of mitotoxic metabolites (BCAA dysmetabolism model).^5^ Although an earlier MR study found no association between BCAAs and FI, our study suggests an inverse association for isoleucine and leucine.^43^ Such inconsistency in findings might be explained by the use of smaller sample size and insufficient considerations on potential pleiotropy when constructing BCAAs instruments in the MR analysis.^43^ Conversely, other MR studies suggested insulin resistance leads to elevated BCAAs.^44^ Additional studies, such as studying metabolic profiles of T2D risk variants related to BCAAs (e.g. *PPM1K*) or interactions with lipids,^5 45^ would help disentangle the etiologic role of BCAAs in T2D risk.

The inverse association between tyrosine and T2D risk was inconsistent with earlier observational studies,^35 41^ but in line with another MR study.^8^ According to observational study, tyrosine is associated with reduced insulin sensitivity and insulin secretion,^2^ and hence contributes to increased T2D risk. However, our MR study suggested little evidence for an association of tyrosine in insulin. This is somewhat also consistent with one observational study showing little impact on the association of tyrosine in T2D upon adjustment for insulin despite the reported positive association with T2D risk.^41^ Nevertheless, these inconsistent findings may either imply confounding and biases in earlier studies, or issues with external validity to humans from animal studies as commented elsewhere.^46^ Having said that, animal studies may help identify possible pathways, such as postulated dopamine pathway in animal study,^47^ to be tested and may inform the mechanisms concerning tyrosine in T2D risk and reconcile the discrepancies between observational and MR studies.

From the public health perspective, our study provides potentially more credible evidence that alanine and isoleucine may increase T2D risk. Although it remains unclear whether increase in circulating amino acids is the consequence of high amino acid intake or a dysfunction in amino acid metabolism, or both, a prospective study indicated that BCAAs may partially mediate the association of meat consumption in T2D risk.^48^ Future dietary guidelines and recommendations may look into alanine and isoleucine intake and supplements given their potential to increase T2D risk. However, since our study focused on the general population, more research is needed to explore any potential effect modification by baseline characteristics, such as individuals with BCAA deficiency and obese individuals.

Although our study is less vulnerable to residual confounding than conventional observational studies, there are some limitations. First, MR studies have three main assumptions for valid causal inference. Although we used strong and independent genetic instruments from large GWAS to reduce the risk of weak instrument bias and genetics were less likely to be confounded, violation of exclusion restriction assumption was possible and hence may have explained the discrepancies between the MR and observational studies. However, we excluded instruments exhibiting horizontal pleiotropy by screening the instruments’ association with other NMR metabolomic markers, as well as performing sensitivity analyses which relied on other assumptions and showed similar findings.

Second, we did not use the most updated T2D GWAS from DIAMANTE Consortium as this most updated version did not provide summary statistics without UK Biobank, ^49^ and hence could introduce possible sample overlap between the GWAS of amino acid and T2D and induce biases. Third, we did not investigate sex specific effects due to the use of summary statistics. As such, follow up studies using individual level data can be conducted to explore the shape of the associations for better targeted interventions, such as sex specific dietary guidelines. Lastly, we only used data derived from European populations. Whether these associations apply to other populations, such as Asians, require further investigations.

## Conclusion

Overall, this study highlighted the causal role of alanine, and potentially, isoleucine in increased T2D risk, which may help inform corresponding dietary recommendations. The paradoxical inverse association of tyrosine with T2D risk requires further investigation.

## Data Availability

The summary statistics of NMR metabolites are available on IEU GWAS database (https://gwas.mrcieu.ac.uk/datasets/?gwas_id__icontains=met-d). The summary statistics for T2D of DIAMANTE consortium and FinnGen study are available on (https://diagram-consortium.org/downloads.html) and (https://risteys.finregistry.fi/endpoints/E4_DM2), respectively. The summary statistics of glycemic traits are available from the MAGIC (https://www.magicinvestigators.org).

## Acknowledgment

We would like to thank MRC Integrative Epidemiology Unit at the University of Bristol for providing the summary statistics of NMR metabolites on IEU GWAS database, and Dr Carolina Borges for the discussion on the respective data. We thank the participants and investigators of DIAMANTE consortium and FinnGen study for providing the summary statistics for T2D.We thank MAGIC for providing summary statistics for FG, 2hGlu, HbA_1c_ and FI. Part of this research (Calculation of principal components) has been conducted using the UK Biobank Resource (Application number: 14864).

## Availability of data and materials

The summary statistics of NMR metabolites are available on IEU GWAS database (https://gwas.mrcieu.ac.uk/datasets/?gwas_idicontains=met-d). The summary statistics for T2D of DIAMANTE consortium and FinnGen study are available on (https://diagram-consortium.org/downloads.html) and (https://risteys.finregistry.fi/endpoints/E4_DM2), respectively. The summary statistics of glycemic traits are available from the MAGIC (https://www.magicinvestigators.org).

## Authors’ contributions

SLAY conceptualized the study. JMYM conducted the analysis with feedback from SLAY. YL cross checked the analyses. SL calculated the PCs required to correct for multiple testing using UK Biobank data. JMYM and SLAY interpreted the results. JMYM wrote the first draft of the manuscript with critical feedback and revisions from BH, TWHT, SL, KL, JCYL, and SLAY. All authors gave final approval of the version to be published. JMYM is the guarantor.

---

**Supplementary Figure 1:**
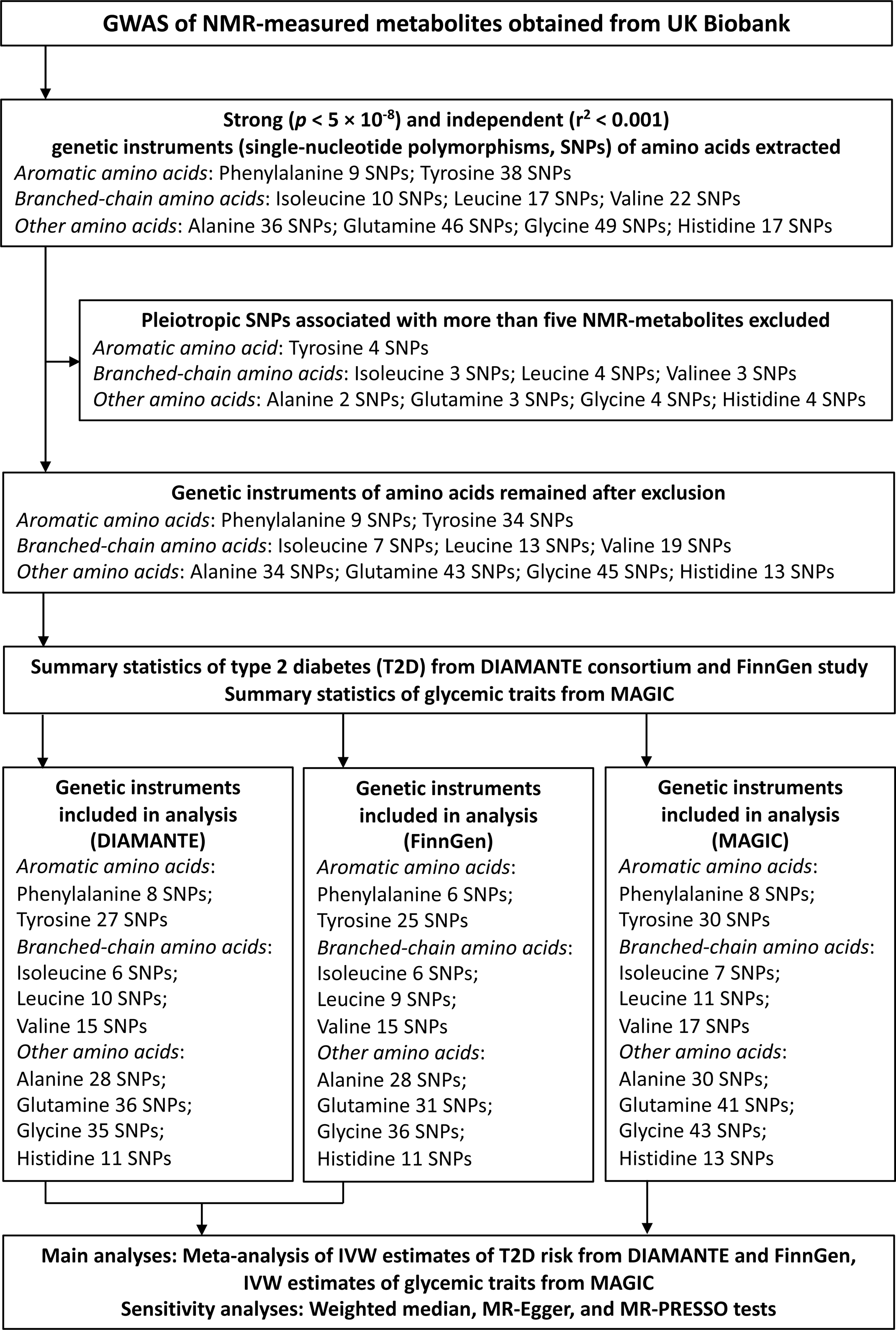
Summary of study design

