## Supplementary Tables for "Evaluating the role of amino acids in type 2 diabetes risk: a Mendelian randomization study"

Spplementary Table 1: Amino acids exposure in this analysis
Spplementary Table 2: Genetic instruments selected for each amino acid exposure
Spplementary Table 3: Genetic instruments excluded with associations with more than five metabolites
Spplementary Table 4: Meta-analysed IVW results of genetically predicted amino acids (per SD) on T2D risk
Spplementary Table 5: Effect of genetically predicted amino acids (per SD) on T2D risk (DIAMANTE, 2018 without UKB)
Spplementary Table 6: Effect of genetically predicted amino acids (per SD) on T2D risk (FinnGen, Round 9)
Spplementary Table 7: Effect of genetically predicted amino acids (per SD) on fasting insulin (log transformed)
Spplementary Table 8: Effect of genetically predicted amino acids (per SD) on HbA1c (%)
Spplementary Table 9: Effect of genetically predicted amino acids (per SD) on fasting glucose (mmol/L)
Spplementary Table 10: Effect of genetically predicted amino acids (per SD) on 2-hour glucose (mmol/L)

Spplementary Table 1: Amino acids exposure in this analysis

| Title | Subgroup | Unit | Sample Size | IEU GWAS ID |
|---|---|---|---|---|
| Alanine | Other amino acid | mmol/L | 115074 | met-d-Ala |
| Glutamine | Other amino acid | mmol/L | 114750 | met-d-Gln |
| Glycine | Other amino acid | mmol/L | 114972 | met-d-Gly |
| Histidine | Other amino acid | mmol/L | 114895 | met-d-His |
| Phenylalanine | Aromatic amino acid | mmol/L | 115025 | met-d-Phe |
| Tyrosine | Aromatic amino acid | mmol/L | 114911 | met-d-Tyr |
| Isoleucine | Branched-chain amino acid | mmol/L | 115075 | met-d-Ile |
| Leucine | Branched-chain amino acid | mmol/L | 115074 | met-d-Leu |
| Valine | Branched-chain amino acid | mmol/L | 115048 | met-d-Val |

Spplementary Table 2: Genetic instruments selected for each amino acid exposure

| Exposure | Subgroup | SNP | Chromosome | Position | Effect allele | Other allele | Effect allele frequency | Beta | Standard error | p-value | Sample size | No. associati | F statistic | $R^2$ | DIAMANTE | FInnGen R9 | Fasting insulin | HbA1c | Fasting glucose | 2-hour glucose |
|---|---|---|---|---|---|---|---|---|---|---|---|---|---|---|---|---|---|---|---|---|
| Alanine | Other | rs79687284 | 1 | 214150821 | C | G | 0.03 | 0.134 | 0.011 | 4.70E-33 | 115074 | 4 | 143 | 1.24E-03 | ✓ | ✓ | ✓ | ✓ | ✓ | ✓ |
| Alanine | Other | rs2160387 | 2 | 65220910 | C | T | 0.43 | -0.045 | 0.004 | 5.90E-28 | 115074 | 1 | 120 | 1.04E-03 | ✓ | ✓ | ✓ | ✓ | ✓ | ✓ |
| Alanine | Other | rs6752053 | 2 | 65666674 | C | T | 0.41 | -0.026 | 0.004 | 5.70E-10 | 115074 | 1 | 38 | 3.34E-04 | ✓ | ✓ | ✓ | ✓ | ✓ | ✓ |
| Alanine | Other | rs11715633 | 3 | 123129946 | A | G | 0.27 | -0.029 | 0.005 | 3.30E-10 | 115074 | 1 | 39 | 3.43E-04 | ✓ | ✓ | ✓ | ✓ | ✓ | ✓ |
| Alanine | Other | rs7624902 | 3 | 160004026 | G | A | 0.54 | -0.039 | 0.004 | 4.10E-21 | 115074 | 1 | 89 | 7.72E-04 | ✓ | ✓ | ✓ | ✓ | ✓ | ✓ |
| Alanine | Other | rs4693210 | 4 | 89208565 | G | A | 0.44 | -0.033 | 0.004 | 1.80E-15 | 115074 | 1 | 63 | 5.49E-04 | ✓ | ✓ | ✓ | ✓ | ✓ | ✓ |
| Alanine | Other | rs138373837 | 5 | 36219710 | T | C | 0.02 | -0.096 | 0.013 | 1.30E-12 | 115074 | 2 | 50 | 4.37E-04 | ✓ | ✓ | ✓ | ✓ | ✓ | ✓ |
| Alanine | Other | rs6931514 | 6 | 20703952 | G | A | 0.26 | 0.037 | 0.005 | 1.40E-15 | 115074 | 1 | 64 | 5.54E-04 | ✓ | ✓ | ✓ | ✓ | ✓ | ✓ |
| Alanine | Other | rs144262262 | 7 | 95222201 | C | T | 0.00 | -0.194 | 0.035 | 2.10E-08 | 115074 | 1 | 31 | 2.73E-04 | ✓ | ✓ | ✓ | ✓ | ✓ | ✓ |
| Alanine | Other | rs2385127 | 8 | 49975746 | A | G | 0.62 | -0.028 | 0.004 | 1.80E-11 | 115074 | 1 | 45 | 3.93E-04 | ✓ | ✓ | ✓ | ✓ | ✓ | ✓ |
| Alanine | Other | rs11558471 | 8 | 118185733 | G | A | 0.31 | -0.031 | 0.004 | 3.10E-12 | 115074 | 1 | 49 | 4.22E-04 | ✓ | ✓ | ✓ | ✓ | ✓ | ✓ |
| Alanine | Other | rs13277542 | 8 | 145747920 | G | T | 0.49 | -0.033 | 0.004 | 4.90E-15 | 115074 | 1 | 61 | 5.32E-04 | NA | ✓ | ✓ | ✓ | ✓ | ✓ |
| Alanine | Other | rs56335308 | 8 | 17419461 | A | G | 0.03 | 0.095 | 0.013 | 3.70E-14 | 115074 | 2 | 57 | 4.98E-04 | ✓ | ✓ | ✓ | ✓ | ✓ | ✓ |
| Alanine | Other | rs12379111 | 9 | 22128180 | G | C | 0.09 | -0.042 | 0.007 | 2.50E-09 | 115074 | 1 | 36 | 3.08E-04 | ✓ | ✓ | ✓ | ✓ | ✓ | ✓ |
| Alanine | Other | rs4237150 | 9 | 4290085 | C | G | 0.40 | 0.028 | 0.004 | 9.10E-12 | 115074 | 2 | 47 | 4.04E-04 | ✓ | NA | NA | NA | NA | NA |
| Alanine | Other | rs2168101 | 11 | 8255408 | A | C | 0.31 | -0.053 | 0.005 | 9.00E-31 | 115074 | 5 | 133 | 1.15E-03 | ✓ | ✓ | ✓ | ✓ | ✓ | ✓ |
| Alanine | Other | rs7925445 | 11 | 18398958 | G | A | 0.56 | 0.030 | 0.004 | 5.60E-13 | 115074 | 2 | 52 | 4.52E-04 | ✓ | NA | ✓ | ✓ | ✓ | ✓ |
| Alanine | Other | rs74093304 | 12 | 59923805 | A | G | 0.12 | 0.050 | 0.006 | 2.30E-15 | 115074 | 1 | 63 | 5.46E-04 | ✓ | ✓ | ✓ | ✓ | ✓ | ✓ |
| Alanine | Other | rs2933243 | 12 | 56860577 | A | G | 0.18 | 0.084 | 0.005 | 3.30E-56 | 115074 | 2 | 250 | 2.16E-03 | ✓ | ✓ | ✓ | ✓ | ✓ | ✓ |
| Alanine | Other | rs7978353 | 12 | 122617989 | G | A | 0.40 | 0.028 | 0.004 | 2.00E-11 | 115074 | 1 | 45 | 3.91E-04 | ✓ | ✓ | ✓ | ✓ | ✓ | ✓ |
| Alanine | Other | rs11183581 | 12 | 47122774 | A | C | 0.21 | 0.058 | 0.005 | 1.30E-31 | 115074 | 1 | 137 | 1.19E-03 | ✓ | ✓ | ✓ | ✓ | ✓ | ✓ |
| Alanine | Other | rs4554975 | 12 | 47201814 | G | A | 0.56 | -0.045 | 0.004 | 3.30E-27 | 115074 | 1 | 117 | 1.01E-03 | ✓ | ✓ | ✓ | ✓ | ✓ | ✓ |
| Alanine | Other | rs4766214 | 12 | 4290249 | G | A | 0.46 | -0.025 | 0.004 | 8.30E-10 | 115074 | 2 | 38 | 3.27E-04 | ✓ | ✓ | ✓ | ✓ | ✓ | ✓ |
| Alanine | Other | rs5023078 | 12 | 133142118 | C | T | 0.53 | -0.024 | 0.004 | 2.80E-09 | 115074 | 1 | 35 | 3.07E-04 | NA | NA | ✓ | ✓ | ✓ | ✓ |
| Alanine | Other | rs58116791 | 14 | 56680822 | C | T | 0.23 | 0.027 | 0.005 | 4.20E-08 | 115074 | 1 | 30 | 2.61E-04 | ✓ | ✓ | ✓ | ✓ | ✓ | ✓ |
| Alanine | Other | rs339969 | 15 | 60883281 | A | C | 0.62 | 0.024 | 0.004 | 7.30E-09 | 115074 | 1 | 33 | 2.91E-04 | ✓ | ✓ | ✓ | ✓ | ✓ | ✓ |
| Alanine | Other | rs8056893 | 16 | 68304392 | A | C | 0.73 | -0.026 | 0.005 | 2.80E-08 | 115074 | 1 | 31 | 2.68E-04 | ✓ | ✓ | ✓ | ✓ | ✓ | ✓ |
| Alanine | Other | rs34495668 | 16 | 70658300 | C | T | 0.10 | 0.054 | 0.007 | 1.10E-14 | 115074 | 1 | 60 | 5.18E-04 | ✓ | ✓ | ✓ | ✓ | ✓ | ✓ |
| Alanine | Other | rs8061221 | 16 | 70078960 | A | G | 0.75 | 0.058 | 0.005 | 4.00E-35 | 115074 | 1 | 153 | 1.33E-03 | NA | ✓ | ✓ | ✓ | ✓ | ✓ |
| Alanine | Other | rs7503139 | 17 | 79641534 | T | C | 0.17 | -0.041 | 0.005 | 1.30E-13 | 115074 | 1 | 55 | 4.77E-04 | ✓ | ✓ | ✓ | ✓ | ✓ | ✓ |
| Alanine | Other | rs73201506 | 21 | 33044408 | G | A | 0.05 | 0.056 | 0.010 | 8.60E-09 | 115074 | 1 | 33 | 2.88E-04 | ✓ | ✓ | ✓ | ✓ | ✓ | ✓ |
| Glutamine | Other | rs79687284 | 1 | 214150821 | C | G | 0.03 | 0.154 | 0.011 | 1.30E-43 | 114750 | 4 | 185 | 1.61E-03 | ✓ | ✓ | ✓ | ✓ | ✓ | ✓ |
| Glutamine | Other | rs13396296 | 2 | 211367095 | C | A | 0.44 | -0.025 | 0.004 | 4.90E-09 | 114750 | 1 | 32 | 2.77E-04 | NA | NA | ✓ | ✓ | ✓ | ✓ |
| Glutamine | Other | rs838737 | 2 | 234325052 | A | G | 0.57 | -0.024 | 0.004 | 2.10E-08 | 114750 | 1 | 32 | 2.80E-04 | ✓ | ✓ | ✓ | ✓ | ✓ | ✓ |
| Glutamine | Other | rs62182473 | 2 | 191740996 | T | C | 0.27 | -0.071 | 0.005 | 8.10E-52 | 114750 | 1 | 227 | 1.97E-03 | ✓ | ✓ | ✓ | ✓ | ✓ | ✓ |
| Glutamine | Other | rs1274961 | 3 | 39191335 | C | T | 0.78 | -0.029 | 0.005 | 4.10E-09 | 114750 | 1 | 34 | 2.96E-04 | ✓ | ✓ | ✓ | ✓ | ✓ | ✓ |
| Glutamine | Other | rs3851365 | 3 | 160191236 | A | C | 0.48 | -0.042 | 0.004 | 1.10E-24 | 114750 | 1 | 102 | 8.89E-04 | ✓ | ✓ | ✓ | ✓ | ✓ | ✓ |
| Glutamine | Other | rs13094915 | 3 | 52507719 | C | G | 0.37 | 0.024 | 0.004 | 2.00E-08 | 114750 | 1 | 31 | 2.72E-04 | ✓ | NA | NA | NA | NA | NA |
| Glutamine | Other | rs73186030 | 3 | 122013465 | T | C | 0.13 | 0.032 | 0.006 | 3.10E-08 | 114750 | 1 | 28 | 2.43E-04 | ✓ | ✓ | ✓ | ✓ | ✓ | ✓ |
| Glutamine | Other | rs3988 | 4 | 18241903 | G | A | 0.40 | -0.022 | 0.004 | 1.50E-08 | 114750 | 1 | 28 | 2.44E-04 | ✓ | ✓ | ✓ | ✓ | ✓ | ✓ |
| Glutamine | Other | rs9687846 | 5 | 55861894 | A | G | 0.20 | -0.029 | 0.005 | 4.30E-08 | 114750 | 4 | 32 | 2.74E-04 | ✓ | ✓ | ✓ | ✓ | ✓ | ✓ |
| Glutamine | Other | rs147139486 | 5 | 128218645 | A | G | 0.05 | -0.051 | 0.009 | 3.40E-08 | 114750 | 1 | 31 | 2.67E-04 | ✓ | ✓ | ✓ | ✓ | ✓ | ✓ |
| Glutamine | Other | rs35261542 | 6 | 20675792 | A | C | 0.26 | 0.030 | 0.005 | 2.90E-10 | 114750 | 1 | 40 | 3.45E-04 | ✓ | ✓ | ✓ | ✓ | ✓ | ✓ |
| Glutamine | Other | rs9381075 | 6 | 41515224 | A | G | 0.26 | 0.025 | 0.005 | 3.20E-08 | 114750 | 1 | 29 | 2.54E-04 | ✓ | ✓ | ✓ | ✓ | ✓ | ✓ |
| Glutamine | Other | rs9482770 | 6 | 127443092 | C | T | 0.45 | -0.033 | 0.004 | 2.20E-15 | 114750 | 1 | 63 | 5.49E-04 | ✓ | ✓ | ✓ | ✓ | ✓ | ✓ |
| Glutamine | Other | rs11422720 | 6 | 131877966 | G | GC | 0.78 | 0.036 | 0.005 | 2.40E-13 | 114750 | 1 | 52 | 4.54E-04 | NA | NA | ✓ | ✓ | ✓ | ✓ |
| Glutamine | Other | rs13234131 | 7 | 73025975 | G | A | 0.13 | 0.072 | 0.006 | 2.60E-32 | 114750 | 3 | 138 | 1.20E-03 | ✓ | ✓ | ✓ | ✓ | ✓ | ✓ |
| Glutamine | Other | rs58673065 | 7 | 1885600 | G | A | 0.23 | 0.029 | 0.005 | 1.90E-09 | 114750 | 1 | 36 | 3.11E-04 | ✓ | ✓ | ✓ | ✓ | ✓ | ✓ |
| Glutamine | Other | rs56335308 | 8 | 17419461 | A | G | 0.03 | 0.116 | 0.013 | 4.90E-20 | 114750 | 2 | 83 | 7.27E-04 | ✓ | ✓ | ✓ | ✓ | ✓ | ✓ |
| Glutamine | Other | rs4237150 | 9 | 4290085 | C | G | 0.40 | 0.033 | 0.004 | 3.90E-15 | 114750 | 2 | 63 | 5.48E-04 | ✓ | NA | NA | NA | NA | NA |
| Glutamine | Other | rs10811663 | 9 | 22139220 | A | G | 0.09 | -0.047 | 0.008 | 1.10E-08 | 114750 | 1 | 34 | 2.97E-04 | ✓ | ✓ | ✓ | ✓ | ✓ | ✓ |
| Glutamine | Other | rs7078003 | 10 | 99359412 | T | C | 0.18 | 0.104 | 0.005 | 6.50E-82 | 114750 | 1 | 369 | 3.21E-03 | ✓ | ✓ | ✓ | ✓ | ✓ | ✓ |
| Glutamine | Other | rs17096421 | 10 | 88820592 | T | A | 0.06 | -0.052 | 0.009 | 3.30E-09 | 114750 | 3 | 33 | 2.90E-04 | ✓ | ✓ | ✓ | ✓ | ✓ | ✓ |
| Glutamine | Other | rs4128598 | 10 | 114785939 | G | A | 0.47 | -0.024 | 0.004 | 4.90E-09 | 114750 | 1 | 34 | 2.95E-04 | ✓ | NA | ✓ | ✓ | ✓ | ✓ |
| Glutamine | Other | rs1750768 | 10 | 22926227 | T | C | 0.04 | 0.058 | 0.010 | 1.40E-08 | 114750 | 1 | 32 | 2.76E-04 | ✓ | ✓ | ✓ | ✓ | ✓ | ✓ |
| Glutamine | Other | rs7925445 | 11 | 18398958 | G | A | 0.56 | 0.028 | 0.004 | 1.50E-11 | 114750 | 2 | 46 | 4.02E-04 | ✓ | NA | ✓ | ✓ | ✓ | ✓ |
| Glutamine | Other | rs2168101 | 11 | 8255408 | A | C | 0.31 | -0.067 | 0.005 | 7.60E-48 | 114750 | 5 | 212 | 1.84E-03 | ✓ | ✓ | ✓ | ✓ | ✓ | ✓ |
| Glutamine | Other | rs78431863 | 11 | 74109553 | T | C | 0.04 | 0.076 | 0.010 | 1.90E-13 | 114750 | 1 | 53 | 4.59E-04 | ✓ | NA | ✓ | ✓ | ✓ | ✓ |
| Glutamine | Other | rs2657879 | 12 | 56865338 | G | A | 0.18 | -0.251 | 0.005 | 1.00E-200 | 114750 | 2 | 2209 | 1.89E-02 | ✓ | ✓ | ✓ | ✓ | ✓ | ✓ |
| Glutamine | Other | rs113674212 | 12 | 47198856 | A | C | 0.07 | 0.135 | 0.008 | 1.70E-60 | 114750 | 1 | 271 | 2.35E-03 | ✓ | ✓ | ✓ | ✓ | ✓ | ✓ |
| Glutamine | Other | rs35752457 | 12 | 59945864 | A | G | 0.16 | 0.066 | 0.006 | 5.40E-29 | 114750 | 1 | 124 | 1.08E-03 | ✓ | NA | ✓ | ✓ | ✓ | ✓ |
| Glutamine | Other | rs4766214 | 12 | 4290249 | G | A | 0.46 | -0.029 | 0.004 | 5.30E-12 | 114750 | 2 | 47 | 4.13E-04 | ✓ | ✓ | ✓ | ✓ | ✓ | ✓ |
| Glutamine | Other | rs4365129 | 12 | 47229840 | T | A | 0.63 | -0.027 | 0.004 | 2.90E-10 | 114750 | 1 | 39 | 3.42E-04 | ✓ | ✓ | ✓ | ✓ | ✓ | ✓ |
| Glutamine | Other | rs7966322 | 12 | 121401846 | T | C | 0.54 | -0.025 | 0.004 | 1.10E-08 | 114750 | 1 | 32 | 2.80E-04 | NA | NA | ✓ | ✓ | ✓ | ✓ |
| Glutamine | Other | rs142525555 | 12 | 57232374 | A | AT | 0.95 | 0.061 | 0.010 | 1.40E-10 | 114750 | 1 | 39 | 3.40E-04 | NA | ✓ | ✓ | ✓ | ✓ | ✓ |
| Glutamine | Other | rs1998848 | 14 | 21492229 | A | G | 0.01 | 0.167 | 0.021 | 1.60E-16 | 114750 | 1 | 64 | 5.54E-04 | ✓ | ✓ | ✓ | ✓ | ✓ | ✓ |
| Glutamine | Other | rs35007880 | 14 | 100795139 | T | G | 0.51 | 0.032 | 0.004 | 5.50E-15 | 114750 | 1 | 61 | 5.27E-04 | ✓ | ✓ | ✓ | ✓ | ✓ | ✓ |
| Glutamine | Other | rs28929474 | 14 | 94844947 | T | C | 0.02 | 0.091 | 0.015 | 6.60E-10 | 114750 | 4 | 37 | 3.26E-04 | ✓ | ✓ | ✓ | ✓ | ✓ | ✓ |
| Glutamine | Other | rs7147721 | 14 | 75186010 | G | A | 0.46 | 0.025 | 0.004 | 7.60E-10 | 114750 | 1 | 38 | 3.27E-04 | NA | NA | ✓ | ✓ | ✓ | ✓ |
| Glutamine | Other | rs35259849 | 16 | 70308837 | T | TA | 0.88 | 0.037 | 0.006 | 2.70E-09 | 114750 | 1 | 34 | 2.93E-04 | NA | NA | ✓ | ✓ | ✓ | ✓ |
| Glutamine | Other | rs8074974 | 17 | 25595818 | C | T | 0.45 | 0.027 | 0.004 | 1.30E-10 | 114750 | 1 | 43 | 3.72E-04 | ✓ | ✓ | ✓ | ✓ | ✓ | ✓ |
| Glutamine | Other | rs11397613 | 18 | 31556350 | TG | T | 0.43 | -0.023 | 0.004 | 1.70E-08 | 114750 | 1 | 30 | 2.64E-04 | NA | ✓ | ✓ | ✓ | ✓ | ✓ |
| Glutamine | Other | rs2039098 | 20 | 56112882 | T | C | 0.60 | -0.024 | 0.004 | 4.50E-09 | 114750 | 1 | 33 | 2.90E-04 | ✓ | NA | ✓ | ✓ | ✓ | ✓ |
| Glutamine | Other | rs738408 | 22 | 44324730 | T | C | 0.22 | 0.033 | 0.005 | 3.50E-11 | 114750 | 1 | 44 | 3.83E-04 | ✓ | ✓ | ✓ | ✓ | ✓ | ✓ |
| Glycine | Other | rs7515955 | 1 | 76143884 | T | A | 0.33 | 0.042 | 0.004 | 3.30E-26 | 114972 | 1 | 102 | 8.84E-04 | NA | ✓ | ✓ | ✓ | ✓ | ✓ |
| Glycine | Other | rs79687284 | 1 | 214150821 | C | G | 0.03 | 0.068 | 0.011 | 7.80E-11 | 114972 | 4 | 41 | 3.52E-04 | ✓ | ✓ | ✓ | ✓ | ✓ | ✓ |
| Glycine | Other | rs6587644 | 1 | 151994458 | A | G | 0.30 | 0.022 | 0.004 | 2.40E-08 | 114972 | 1 | 27 | 2.34E-04 | ✓ | ✓ | ✓ | ✓ | ✓ | ✓ |
| Glycine | Other | rs1801133 | 1 | 11856378 | A | G | 0.34 | 0.023 | 0.004 | 1.80E-08 | 114972 | 1 | 32 | 2.79E-04 | ✓ | ✓ | ✓ | ✓ | ✓ | ✓ |
| Glycine | Other | rs561931 | 1 | 120254506 | G | A | 0.58 | 0.029 | 0.004 | 4.50E-14 | 114972 | 1 | 54 | 4.70E-04 | ✓ | ✓ | ✓ | ✓ | ✓ | ✓ |
| Glycine | Other | rs35034344 | 2 | 211026796 | T | A | 0.27 | -0.065 | 0.005 | 1.90E-46 | 114972 | 1 | 203 | 1.76E-03 | NA | NA | ✓ | ✓ | ✓ | ✓ |
| Glycine | Other | rs4380169 | 2 | 212145768 | C | T | 0.49 | -0.042 | 0.004 | 3.60E-27 | 114972 | 1 | 117 | 1.02E-03 | ✓ | ✓ | ✓ | ✓ | ✓ | ✓ |
| Glycine | Other | rs75604103 | 2 | 211692010 | G | A | 0.12 | -0.099 | 0.006 | 1.10E-60 | 114972 | 1 | 272 | 2.36E-03 | ✓ | ✓ | ✓ | ✓ | ✓ | ✓ |
| Glycine | Other | rs36105243 | 2 | 165524751 | G | C | 0.35 | 0.026 | 0.004 | 5.30E-11 | 114972 | 1 | 40 | 3.50E-04 | ✓ | ✓ | ✓ | ✓ | ✓ | ✓ |
| Glycine | Other | rs10190808 | 2 | 211993631 | C | G | 0.14 | 0.035 | 0.006 | 2.40E-09 | 114972 | 1 | 37 | 3.24E-04 | ✓ | ✓ | ✓ | ✓ | ✓ | ✓ |
| Glycine | Other | rs17722201 | 2 | 209645912 | C | T | 0.22 | 0.028 | 0.005 | 9.80E-09 | 114972 | 1 | 34 | 2.98E-04 | ✓ | NA | ✓ | ✓ | ✓ | ✓ |
| Glycine | Other | rs10197334 | 2 | 211371267 | A | G | 0.12 | 0.037 | 0.006 | 2.20E-09 | 114972 | 1 | 38 | 3.29E-04 | ✓ | ✓ | ✓ | ✓ | ✓ | ✓ |
| Glycine | Other | rs2972154 | 2 | 227065811 | T | C | 0.54 | -0.022 | 0.004 | 3.10E-08 | 114972 | 1 | 33 | 2.89E-04 | ✓ | ✓ | ✓ | ✓ | ✓ | ✓ |
| Glycine | Other | rs62203720 | 2 | 211482273 | G | T | 0.02 | 0.135 | 0.017 | 5.60E-17 | 114972 | 1 | 66 | 5.72E-04 | ✓ | ✓ | ✓ | ✓ | ✓ | ✓ |
| Glycine | Other | rs3836259 | 3 | 125830955 | A | AG | 0.78 | 0.030 | 0.005 | 1.70E-10 | 114972 | 1 | 41 | 3.54E-04 | NA | ✓ | ✓ | ✓ | ✓ | ✓ |
| Glycine | Other | rs10934753 | 3 | 125906179 | A | G | 0.42 | 0.069 | 0.004 | 3.40E-74 | 114972 | 1 | 303 | 2.63E-03 | ✓ | ✓ | ✓ | ✓ | ✓ | ✓ |
| Glycine | Other | rs35408322 | 3 | 12360357 | G | GT | 0.88 | -0.034 | 0.006 | 1.00E-08 | 114972 | 1 | 33 | 2.90E-04 | NA | NA | ✓ | ✓ | ✓ | ✓ |
| Glycine | Other | rs1965869 | 4 | 89677537 | C | T | 0.72 | -0.024 | 0.004 | 9.00E-09 | 114972 | 1 | 31 | 2.72E-04 | ✓ | ✓ | ✓ | ✓ | ✓ | ✓ |
| Glycine | Other | rs11242109 | 5 | 131677047 | T | G | 0.48 | 0.023 | 0.004 | 2.00E-08 | 114972 | 2 | 34 | 2.92E-04 | ✓ | ✓ | ✓ | ✓ | ✓ | ✓ |
| Glycine | Other | rs7704653 | 5 | 90255685 | G | A | 0.72 | -0.032 | 0.004 | 3.70E-13 | 114972 | 1 | 52 | 4.48E-04 | ✓ | ✓ | ✓ | ✓ | ✓ | ✓ |
| Glycine | Other | rs555754 | 6 | 160769423 | A | G | 0.47 | 0.023 | 0.004 | 1.20E-09 | 114972 | 1 | 35 | 3.08E-04 | ✓ | ✓ | ✓ | ✓ | ✓ | ✓ |
| Glycine | Other | rs2608913 | 6 | 131870261 | C | T | 0.22 | -0.024 | 0.005 | 2.30E-08 | 114972 | 1 | 26 | 2.30E-04 | ✓ | ✓ | ✓ | ✓ | ✓ | ✓ |
| Glycine | Other | rs7800001 | 7 | 56072010 | C | T | 0.76 | 0.072 | 0.005 | 5.50E-61 | 114972 | 1 | 253 | 2.19E-03 | ✓ | ✓ | ✓ | ✓ | ✓ | ✓ |
| Glycine | Other | rs192322963 | 8 | 17445955 | A | G | 0.03 | 0.083 | 0.013 | 4.10E-11 | 114972 | 1 | 44 | 3.81E-04 | NA | ✓ | ✓ | ✓ | ✓ | ✓ |
| Glycine | Other | rs6601302 | 8 | 9239458 | G | T | 0.75 | 0.028 | 0.005 | 2.10E-10 | 114972 | 1 | 38 | 3.27E-04 | ✓ | ✓ | ✓ | ✓ | ✓ | ✓ |
| Glycine | Other | rs28435239 | 9 | 5989087 | A | G | 0.77 | -0.029 | 0.005 | 8.60E-10 | 114972 | 1 | 39 | 3.37E-04 | ✓ | ✓ | ✓ | ✓ | ✓ | ✓ |
| Glycine | Other | rs2026972 | 9 | 6538279 | C | G | 0.31 | -0.082 | 0.004 | 2.20E-87 | 114972 | 1 | 377 | 3.27E-03 | ✓ | ✓ | ✓ | ✓ | ✓ | ✓ |
| Glycine | Other | rs2168101 | 11 | 8255408 | A | C | 0.31 | -0.029 | 0.004 | 3.90E-11 | 114972 | 5 | 46 | 3.96E-04 | ✓ | ✓ | ✓ | ✓ | ✓ | ✓ |
| Glycine | Other | rs2657879 | 12 | 56865338 | G | A | 0.18 | 0.048 | 0.005 | 5.20E-22 | 114972 | 2 | 90 | 7.85E-04 | ✓ | ✓ | ✓ | ✓ | ✓ | ✓ |
| Glycine | Other | rs67523949 | 12 | 348506 | T | C | 0.54 | 0.026 | 0.004 | 3.30E-11 | 114972 | 1 | 45 | 3.94E-04 | ✓ | NA | ✓ | ✓ | ✓ | ✓ |
| Glycine | Other | rs11172190 | 12 | 57766305 | T | C | 0.50 | 0.024 | 0.004 | 1.00E-10 | 114972 | 1 | 37 | 3.19E-04 | ✓ | ✓ | ✓ | ✓ | ✓ | ✓ |
| Glycine | Other | rs2711697 | 12 | 47265729 | C | A | 0.37 | 0.027 | 0.004 | 1.10E-11 | 114972 | 1 | 43 | 3.76E-04 | ✓ | ✓ | ✓ | ✓ | ✓ | ✓ |
| Glycine | Other | rs56819961 | 12 | 47137673 | C | T | 0.21 | 0.043 | 0.005 | 1.40E-20 | 114972 | 1 | 80 | 6.96E-04 | ✓ | ✓ | ✓ | ✓ | ✓ | ✓ |
| Glycine | Other | rs11045886 | 12 | 21386493 | C | A | 0.17 | 0.031 | 0.005 | 8.30E-09 | 114972 | 1 | 34 | 2.94E-04 | NA | NA | ✓ | ✓ | ✓ | ✓ |
| Glycine | Other | rs9532939 | 13 | 42440496 | A | T | 0.35 | 0.025 | 0.004 | 8.70E-09 | 114972 | 1 | 35 | 3.04E-04 | NA | NA | ✓ | ✓ | ✓ | ✓ |
| Glycine | Other | rs149181595 | 15 | 43685807 | C | A | 0.03 | 0.094 | 0.012 | 3.90E-17 | 114972 | 1 | 63 | 5.44E-04 | ✓ | ✓ | ✓ | ✓ | ✓ | ✓ |
| Glycine | Other | rs34945403 | 15 | 58430763 | G | A | 0.07 | -0.063 | 0.008 | 1.70E-15 | 114972 | 1 | 62 | 5.42E-04 | ✓ | ✓ | ✓ | ✓ | ✓ | ✓ |
| Glycine | Other | rs4889229 | 16 | 81113672 | T | C | 0.92 | 0.118 | 0.007 | 4.20E-64 | 114972 | 1 | 275 | 2.39E-03 | ✓ | ✓ | ✓ | ✓ | ✓ | ✓ |
| Glycine | Other | rs112247225 | 16 | 81154900 | T | C | 0.05 | -0.128 | 0.009 | 5.20E-45 | 114972 | 1 | 191 | 1.66E-03 | ✓ | ✓ | ✓ | ✓ | ✓ | ✓ |
| Glycine | Other | rs7188156 | 16 | 79938114 | G | T | 0.14 | 0.032 | 0.006 | 1.20E-08 | 114972 | 1 | 33 | 2.83E-04 | ✓ | ✓ | ✓ | ✓ | ✓ | ✓ |
| Glycine | Other | rs369648654 | 17 | 45498226 | T | G | 0.49 | -0.040 | 0.004 | 2.20E-24 | 114972 | 1 | 103 | 8.93E-04 | NA | NA | ✓ | ✓ | ✓ | ✓ |
| Glycine | Other | rs11666281 | 19 | 18234588 | T | C | 0.25 | -0.037 | 0.004 | 1.30E-17 | 114972 | 1 | 69 | 5.96E-04 | ✓ | ✓ | ✓ | ✓ | ✓ | ✓ |
| Glycine | Other | rs56113850 | 19 | 41353107 | C | T | 0.58 | -0.022 | 0.004 | 2.40E-08 | 114972 | 1 | 31 | 2.67E-04 | ✓ | ✓ | ✓ | ✓ | ✓ | ✓ |
| Histidine | Other | rs141050588 | 1 | 169213261 | C | CAATTAAAT | 0.59 | 0.025 | 0.004 | 3.10E-09 | 114895 | 1 | 35 | 3.06E-04 | NA | NA | ✓ | ✓ | ✓ | ✓ |
| Histidine | Other | rs780094 | 2 | 27741237 | C | T | 0.62 | 0.034 | 0.004 | 9.90E-16 | 114895 | 3 | 64 | 5.61E-04 | ✓ | ✓ | ✓ | ✓ | ✓ | ✓ |
| Histidine | Other | rs34004251 | 3 | 148929951 | A | T | 0.18 | -0.054 | 0.005 | 3.50E-23 | 114895 | 1 | 98 | 8.56E-04 | ✓ | ✓ | ✓ | ✓ | ✓ | ✓ |
| Histidine | Other | rs117591301 | 6 | 134872112 | T | A | 0.03 | -0.078 | 0.013 | 1.00E-09 | 114895 | 1 | 37 | 3.24E-04 | ✓ | ✓ | ✓ | ✓ | ✓ | ✓ |
| Histidine | Other | rs2272662 | 8 | 145639726 | C | T | 0.58 | 0.029 | 0.004 | 3.70E-12 | 114895 | 1 | 48 | 4.20E-04 | ✓ | ✓ | ✓ | ✓ | ✓ | ✓ |
| Histidine | Other | rs2168101 | 11 | 8255408 | A | C | 0.31 | -0.027 | 0.005 | 3.50E-09 | 114895 | 5 | 35 | 3.04E-04 | ✓ | ✓ | ✓ | ✓ | ✓ | ✓ |
| Histidine | Other | rs150670499 | 12 | 96442150 | C | CTATG | 0.87 | -0.055 | 0.007 | 1.40E-15 | 114895 | 1 | 64 | 5.55E-04 | NA | NA | ✓ | ✓ | ✓ | ✓ |
| Histidine | Other | rs75341400 | 12 | 96315168 | T | G | 0.10 | -0.088 | 0.007 | 4.00E-36 | 114895 | 1 | 157 | 1.37E-03 | ✓ | ✓ | ✓ | ✓ | ✓ | ✓ |
| Histidine | Other | rs61938133 | 12 | 96120605 | A | G | 0.04 | 0.080 | 0.011 | 7.80E-14 | 114895 | 1 | 56 | 4.86E-04 | ✓ | ✓ | ✓ | ✓ | ✓ | ✓ |
| Histidine | Other | rs76191003 | 12 | 47182531 | T | C | 0.07 | 0.050 | 0.008 | 1.40E-09 | 114895 | 1 | 37 | 3.18E-04 | ✓ | ✓ | ✓ | ✓ | ✓ | ✓ |
| Histidine | Other | rs2939302 | 12 | 56860020 | A | G | 0.18 | 0.059 | 0.005 | 1.10E-28 | 114895 | 1 | 123 | 1.07E-03 | ✓ | ✓ | ✓ | ✓ | ✓ | ✓ |
| Histidine | Other | rs709932 | 14 | 94849201 | T | C | 0.16 | 0.039 | 0.006 | 5.50E-12 | 114895 | 1 | 48 | 4.13E-04 | ✓ | ✓ | ✓ | ✓ | ✓ | ✓ |
| Histidine | Other | rs1958393 | 14 | 21516447 | G | C | 0.11 | 0.061 | 0.007 | 5.90E-20 | 114895 | 1 | 84 | 7.27E-04 | ✓ | ✓ | ✓ | ✓ | ✓ | ✓ |
| Phenylalanine | Aromatic | rs715 | 2 | 211543055 | C | T | 0.31 | -0.028 | 0.004 | 4.20E-10 | 115025 | 3 | 39 | 3.39E-04 | ✓ | ✓ | ✓ | ✓ | ✓ | ✓ |
| Phenylalanine | Aromatic | rs2180314 | 6 | 52617731 | G | C | 0.60 | 0.023 | 0.004 | 4.10E-08 | 115025 | 1 | 30 | 2.62E-04 | ✓ | NA | NA | NA | NA | NA |
| Phenylalanine | Aromatic | rs9467618 | 6 | 25824809 | T | C | 0.26 | -0.030 | 0.005 | 1.70E-10 | 115025 | 1 | 41 | 3.54E-04 | ✓ | ✓ | ✓ | ✓ | ✓ | ✓ |
| Phenylalanine | Aromatic | rs10750864 | 11 | 57259694 | A | T | 0.74 | 0.058 | 0.005 | 5.00E-34 | 115025 | 1 | 148 | 1.28E-03 | NA | NA | ✓ | ✓ | ✓ | ✓ |
| Phenylalanine | Aromatic | rs1498694 | 12 | 103257876 | G | A | 0.61 | -0.109 | 0.004 | 1.60E-146 | 115025 | 1 | 664 | 5.74E-03 | ✓ | ✓ | ✓ | ✓ | ✓ | ✓ |
| Phenylalanine | Aromatic | rs2933235 | 12 | 56921418 | G | A | 0.17 | 0.033 | 0.006 | 4.30E-09 | 115025 | 1 | 34 | 3.00E-04 | ✓ | NA | ✓ | ✓ | ✓ | ✓ |
| Phenylalanine | Aromatic | rs7969789 | 12 | 103325781 | T | C | 0.33 | -0.063 | 0.004 | 4.80E-46 | 115025 | 1 | 203 | 1.76E-03 | ✓ | ✓ | ✓ | ✓ | ✓ | ✓ |
| Phenylalanine | Aromatic | rs17253619 | 14 | 55411744 | C | T | 0.12 | 0.068 | 0.006 | 1.30E-26 | 115025 | 1 | 114 | 9.90E-04 | ✓ | ✓ | ✓ | ✓ | ✓ | ✓ |
| Phenylalanine | Aromatic | rs4781730 | 16 | 16254626 | T | C | 0.28 | -0.026 | 0.005 | 1.90E-08 | 115025 | 1 | 32 | 2.74E-04 | ✓ | ✓ | ✓ | ✓ | ✓ | ✓ |
| Tyrosine | Aromatic | rs715 | 2 | 211543055 | C | T | 0.31 | -0.030 | 0.004 | 9.90E-12 | 114911 | 3 | 46 | 4.03E-04 | ✓ | ✓ | ✓ | ✓ | ✓ | ✓ |
| Tyrosine | Aromatic | rs114232169 | 3 | 186389769 | G | T | 0.67 | -0.032 | 0.004 | 1.90E-13 | 114911 | 1 | 54 | 4.71E-04 | NA | NA | ✓ | ✓ | ✓ | ✓ |
| Tyrosine | Aromatic | rs1433210 | 4 | 124766956 | C | A | 0.25 | -0.027 | 0.005 | 3.00E-08 | 114911 | 1 | 31 | 2.67E-04 | ✓ | ✓ | ✓ | ✓ | ✓ | ✓ |
| Tyrosine | Aromatic | rs1126671 | 4 | 100048414 | C | T | 0.70 | -0.041 | 0.004 | 1.70E-19 | 114911 | 3 | 82 | 7.09E-04 | ✓ | ✓ | ✓ | ✓ | ✓ | ✓ |
| Tyrosine | Aromatic | rs6868618 | 5 | 90191256 | G | A | 0.38 | 0.025 | 0.004 | 3.00E-09 | 114911 | 1 | 35 | 3.06E-04 | ✓ | ✓ | ✓ | ✓ | ✓ | ✓ |
| Tyrosine | Aromatic | rs75661418 | 6 | 111489659 | G | A | 0.02 | -0.082 | 0.013 | 9.70E-10 | 114911 | 1 | 37 | 3.25E-04 | ✓ | ✓ | ✓ | ✓ | ✓ | ✓ |
| Tyrosine | Aromatic | rs35614968 | 6 | 112020241 | A | G | 0.03 | -0.122 | 0.013 | 1.20E-21 | 114911 | 1 | 91 | 7.95E-04 | ✓ | ✓ | ✓ | ✓ | ✓ | ✓ |
| Tyrosine | Aromatic | rs116862171 | 6 | 112145789 | G | A | 0.02 | 0.093 | 0.016 | 1.00E-08 | 114911 | 1 | 33 | 2.86E-04 | ✓ | ✓ | ✓ | ✓ | ✓ | ✓ |
| Tyrosine | Aromatic | rs9267947 | 6 | 32211218 | G | A | 0.48 | 0.023 | 0.004 | 3.60E-08 | 114911 | 1 | 30 | 2.64E-04 | ✓ | NA | ✓ | ✓ | ✓ | ✓ |
| Tyrosine | Aromatic | rs146133919 | 6 | 112130000 | C | T | 0.03 | -0.078 | 0.013 | 3.10E-09 | 114911 | 1 | 35 | 3.05E-04 | ✓ | ✓ | ✓ | ✓ | ✓ | ✓ |
| Tyrosine | Aromatic | rs12206654 | 6 | 111556834 | C | T | 0.15 | 0.188 | 0.006 | 1.00E-200 | 114911 | 1 | 1027 | 8.85E-03 | ✓ | ✓ | ✓ | ✓ | ✓ | ✓ |
| Tyrosine | Aromatic | rs56236906 | 6 | 111991104 | A | G | 0.02 | 0.084 | 0.015 | 9.30E-09 | 114911 | 1 | 33 | 2.87E-04 | ✓ | ✓ | ✓ | ✓ | ✓ | ✓ |
| Tyrosine | Aromatic | rs7786102 | 7 | 25965759 | A | G | 0.72 | -0.026 | 0.005 | 8.10E-09 | 114911 | 1 | 33 | 2.89E-04 | ✓ | ✓ | ✓ | ✓ | ✓ | ✓ |
| Tyrosine | Aromatic | rs740122 | 7 | 28166442 | A | G | 0.22 | -0.037 | 0.005 | 1.10E-13 | 114911 | 1 | 55 | 4.80E-04 | ✓ | ✓ | ✓ | ✓ | ✓ | ✓ |
| Tyrosine | Aromatic | rs7019778 | 9 | 22134651 | C | A | 0.42 | -0.025 | 0.004 | 2.50E-09 | 114911 | 1 | 36 | 3.09E-04 | ✓ | ✓ | ✓ | ✓ | ✓ | ✓ |
| Tyrosine | Aromatic | rs9414801 | 10 | 65149089 | A | G | 0.53 | -0.036 | 0.004 | 5.30E-18 | 114911 | 1 | 75 | 6.50E-04 | NA | NA | ✓ | ✓ | ✓ | ✓ |

| | | | | | | | | | | | | | | | | | | | | |
|---|---|---|---|---|---|---|---|---|---|---|---|---|---|---|---|---|---|---|---|---|
| Tyrosine | Aromatic | rs174547 | 11 | 61570783 | C | T | 0.34 | 0.034 | 0.004 | 5.10E-15 | 114911 | 1 | 61 | 5.32E-04 | ✓ | ✓ | ✓ | ✓ | ✓ | ✓ |
| Tyrosine | Aromatic | rs2168101 | 11 | 8255408 | A | C | 0.31 | -0.031 | 0.005 | 1.60E-11 | 114911 | 5 | 45 | 3.95E-04 | ✓ | ✓ | ✓ | ✓ | ✓ | ✓ |
| Tyrosine | Aromatic | rs36055409 | 12 | 59990603 | T | G | 0.14 | 0.033 | 0.006 | 2.10E-08 | 114911 | 1 | 31 | 2.73E-04 | ✓ | ✓ | ✓ | ✓ | ✓ | ✓ |
| Tyrosine | Aromatic | rs2230681 | 12 | 122326812 | C | T | 0.87 | -0.042 | 0.006 | 1.40E-11 | 114911 | 1 | 46 | 3.97E-04 | ✓ | ✓ | ✓ | ✓ | ✓ | ✓ |
| Tyrosine | Aromatic | rs4149056 | 12 | 21331549 | C | T | 0.15 | -0.060 | 0.006 | 2.70E-25 | 114911 | 1 | 108 | 9.39E-04 | ✓ | ✓ | ✓ | ✓ | ✓ | ✓ |
| Tyrosine | Aromatic | rs79020793 | 12 | 47199549 | A | G | 0.07 | 0.078 | 0.008 | 3.10E-21 | 114911 | 1 | 90 | 7.78E-04 | ✓ | ✓ | ✓ | ✓ | ✓ | ✓ |
| Tyrosine | Aromatic | rs2638315 | 12 | 56865056 | C | G | 0.18 | 0.067 | 0.005 | 1.10E-36 | 114911 | 4 | 160 | 1.39E-03 | ✓ | ✓ | ✓ | ✓ | ✓ | ✓ |
| Tyrosine | Aromatic | rs194749 | 14 | 69273905 | C | T | 0.24 | 0.027 | 0.005 | 4.10E-08 | 114911 | 1 | 30 | 2.62E-04 | ✓ | ✓ | ✓ | ✓ | ✓ | ✓ |
| Tyrosine | Aromatic | rs6575900 | 14 | 102675350 | C | G | 0.20 | -0.033 | 0.005 | 2.00E-10 | 114911 | 1 | 40 | 3.52E-04 | ✓ | ✓ | ✓ | ✓ | ✓ | ✓ |
| Tyrosine | Aromatic | rs4788811 | 16 | 71625209 | G | A | 0.14 | -0.040 | 0.006 | 2.10E-11 | 114911 | 1 | 45 | 3.91E-04 | ✓ | ✓ | ✓ | ✓ | ✓ | ✓ |
| Tyrosine | Aromatic | rs150851429 | 16 | 71625831 | C | G | 0.02 | 0.399 | 0.017 | 1.90E-120 | 114911 | 1 | 545 | 4.72E-03 | ✓ | NA | ✓ | ✓ | ✓ | ✓ |
| Tyrosine | Aromatic | rs76819459 | 16 | 79718917 | G | A | 0.33 | 0.025 | 0.004 | 1.70E-08 | 114911 | 1 | 32 | 2.76E-04 | ✓ | ✓ | ✓ | ✓ | ✓ | ✓ |
| Tyrosine | Aromatic | rs814573 | 19 | 45424351 | T | A | 0.19 | -0.035 | 0.005 | 2.30E-10 | 114911 | 1 | 40 | 3.50E-04 | NA | NA | ✓ | ✓ | ✓ | ✓ |
| Tyrosine | Aromatic | rs61676179 | 22 | 32176825 | A | C | 0.05 | -0.053 | 0.009 | 2.10E-08 | 114911 | 1 | 31 | 2.73E-04 | ✓ | ✓ | ✓ | ✓ | ✓ | ✓ |
| Isoleucine | Branched chain | rs72538440 | 2 | 65225088 | G | GC | 0.58 | -0.024 | 0.004 | 6.60E-09 | 115075 | 4 | 34 | 2.92E-04 | NA | ✓ | ✓ | ✓ | ✓ | ✓ |
| Isoleucine | Branched chain | rs7656569 | 4 | 89166761 | A | C | 0.19 | 0.046 | 0.005 | 1.40E-18 | 115075 | 4 | 77 | 6.72E-04 | ✓ | ✓ | ✓ | ✓ | ✓ | ✓ |
| Isoleucine | Branched chain | rs10018448 | 4 | 89225171 | G | A | 0.54 | 0.067 | 0.004 | 4.10E-60 | 115075 | 4 | 267 | 2.32E-03 | ✓ | NA | ✓ | ✓ | ✓ | ✓ |
| Isoleucine | Branched chain | rs2941456 | 8 | 76443463 | A | G | 0.20 | 0.029 | 0.005 | 9.10E-09 | 115075 | 1 | 33 | 2.87E-04 | ✓ | ✓ | ✓ | ✓ | ✓ | ✓ |
| Isoleucine | Branched chain | rs7302925 | 12 | 56861458 | G | A | 0.80 | -0.041 | 0.005 | 3.60E-16 | 115075 | 1 | 66 | 5.77E-04 | ✓ | ✓ | ✓ | ✓ | ✓ | ✓ |
| Isoleucine | Branched chain | rs12325419 | 16 | 70368909 | A | G | 0.12 | -0.074 | 0.006 | 1.10E-31 | 115075 | 1 | 137 | 1.19E-03 | ✓ | ✓ | ✓ | ✓ | ✓ | ✓ |
| Isoleucine | Branched chain | rs545587 | 19 | 49319664 | C | A | 0.51 | 0.037 | 0.004 | 2.30E-19 | 115075 | 1 | 81 | 7.03E-04 | ✓ | ✓ | ✓ | ✓ | ✓ | ✓ |
| Leucine | Branched chain | rs11166420 | 1 | 100702216 | A | T | 0.90 | -0.038 | 0.007 | 1.70E-08 | 115074 | 1 | 32 | 2.76E-04 | ✓ | ✓ | ✓ | ✓ | ✓ | ✓ |
| Leucine | Branched chain | rs72538440 | 2 | 65225088 | G | GC | 0.58 | -0.027 | 0.004 | 4.30E-11 | 115074 | 4 | 43 | 3.78E-04 | NA | ✓ | ✓ | ✓ | ✓ | ✓ |
| Leucine | Branched chain | rs150277164 | 4 | 89258580 | A | G | 0.01 | 0.116 | 0.017 | 5.20E-12 | 115074 | 1 | 48 | 4.14E-04 | ✓ | ✓ | ✓ | ✓ | ✓ | ✓ |
| Leucine | Branched chain | rs10018448 | 4 | 89225171 | G | A | 0.54 | 0.088 | 0.004 | 4.30E-108 | 115074 | 4 | 488 | 4.22E-03 | ✓ | NA | ✓ | ✓ | ✓ | ✓ |
| Leucine | Branched chain | rs7656569 | 4 | 89166761 | A | C | 0.19 | 0.056 | 0.005 | 4.20E-28 | 115074 | 4 | 121 | 1.05E-03 | ✓ | ✓ | ✓ | ✓ | ✓ | ✓ |
| Leucine | Branched chain | rs2977929 | 8 | 76454025 | T | C | 0.20 | 0.028 | 0.005 | 1.80E-08 | 115074 | 2 | 32 | 2.76E-04 | ✓ | ✓ | ✓ | ✓ | ✓ | ✓ |
| Leucine | Branched chain | rs35350651 | 12 | 111907431 | AC | A | 0.50 | -0.023 | 0.004 | 4.90E-09 | 115074 | 1 | 34 | 2.97E-04 | NA | ✓ | NA | NA | NA | NA |
| Leucine | Branched chain | rs2638315 | 12 | 56865056 | C | G | 0.18 | 0.041 | 0.005 | 1.20E-15 | 115074 | 4 | 64 | 5.56E-04 | ✓ | ✓ | ✓ | ✓ | ✓ | ✓ |
| Leucine | Branched chain | rs9930957 | 16 | 72149923 | T | C | 0.16 | 0.053 | 0.005 | 2.10E-22 | 115074 | 2 | 95 | 8.23E-04 | ✓ | ✓ | ✓ | ✓ | ✓ | ✓ |
| Leucine | Branched chain | rs12974412 | 19 | 14151809 | G | A | 0.78 | -0.029 | 0.005 | 3.30E-09 | 115074 | 3 | 35 | 3.04E-04 | ✓ | ✓ | ✓ | ✓ | ✓ | ✓ |
| Leucine | Branched chain | rs4801776 | 19 | 49304215 | T | C | 0.30 | -0.032 | 0.004 | 2.40E-13 | 115074 | 2 | 54 | 4.66E-04 | ✓ | NA | ✓ | ✓ | ✓ | ✓ |
| Leucine | Branched chain | rs5747934 | 22 | 18915282 | T | C | 0.04 | -0.057 | 0.010 | 8.60E-09 | 115074 | 2 | 33 | 2.88E-04 | ✓ | NA | ✓ | ✓ | ✓ | ✓ |
| Valine | Branched chain | rs72538440 | 2 | 65225088 | G | GC | 0.58 | -0.064 | 0.004 | 6.20E-55 | 115048 | 4 | 244 | 2.11E-03 | NA | ✓ | ✓ | ✓ | ✓ | ✓ |
| Valine | Branched chain | rs2943652 | 2 | 227108446 | T | C | 0.64 | 0.024 | 0.004 | 1.20E-08 | 115048 | 4 | 32 | 2.82E-04 | ✓ | ✓ | ✓ | ✓ | ✓ | ✓ |
| Valine | Branched chain | rs10018448 | 4 | 89225171 | G | A | 0.54 | 0.105 | 0.004 | 2.30E-148 | 115048 | 4 | 673 | 5.82E-03 | ✓ | NA | ✓ | ✓ | ✓ | ✓ |
| Valine | Branched chain | rs12648408 | 4 | 89244131 | A | G | 0.05 | -0.051 | 0.009 | 4.20E-08 | 115048 | 1 | 30 | 2.61E-04 | NA | ✓ | ✓ | ✓ | ✓ | ✓ |
| Valine | Branched chain | rs7656569 | 4 | 89166761 | A | C | 0.19 | 0.071 | 0.005 | 3.80E-43 | 115048 | 4 | 190 | 1.65E-03 | ✓ | ✓ | ✓ | ✓ | ✓ | ✓ |
| Valine | Branched chain | rs61587941 | 5 | 55814890 | A | G | 0.05 | -0.050 | 0.009 | 3.40E-08 | 115048 | 1 | 30 | 2.65E-04 | ✓ | ✓ | ✓ | ✓ | ✓ | ✓ |
| Valine | Branched chain | rs6941263 | 6 | 109162094 | A | T | 0.19 | -0.029 | 0.005 | 3.40E-08 | 115048 | 1 | 30 | 2.65E-04 | ✓ | ✓ | ✓ | ✓ | ✓ | ✓ |
| Valine | Branched chain | rs17096421 | 10 | 88820592 | T | A | 0.06 | 0.063 | 0.009 | 1.40E-12 | 115048 | 3 | 50 | 4.36E-04 | ✓ | ✓ | ✓ | ✓ | ✓ | ✓ |
| Valine | Branched chain | rs36181536 | 12 | 122528263 | C | T | 0.67 | -0.032 | 0.004 | 1.30E-13 | 115048 | 2 | 55 | 4.76E-04 | ✓ | ✓ | ✓ | ✓ | ✓ | ✓ |
| Valine | Branched chain | rs2638315 | 12 | 56865056 | C | G | 0.18 | 0.043 | 0.005 | 2.20E-16 | 115048 | 4 | 67 | 5.85E-04 | ✓ | ✓ | ✓ | ✓ | ✓ | ✓ |
| Valine | Branched chain | rs2274815 | 14 | 102718052 | A | G | 0.21 | 0.028 | 0.005 | 1.80E-08 | 115048 | 1 | 32 | 2.75E-04 | ✓ | ✓ | ✓ | ✓ | ✓ | ✓ |
| Valine | Branched chain | rs8071084 | 17 | 79634162 | G | T | 0.16 | -0.032 | 0.005 | 3.10E-09 | 115048 | 1 | 35 | 3.05E-04 | ✓ | ✓ | ✓ | ✓ | ✓ | ✓ |
| Valine | Branched chain | rs35230038 | 19 | 49300431 | A | G | 0.05 | -0.098 | 0.010 | 1.10E-23 | 115048 | 1 | 101 | 8.74E-04 | ✓ | ✓ | ✓ | ✓ | ✓ | ✓ |
| Valine | Branched chain | rs117048185 | 19 | 49309776 | C | G | 0.02 | 0.190 | 0.015 | 8.70E-35 | 115048 | 1 | 151 | 1.31E-03 | ✓ | ✓ | ✓ | ✓ | ✓ | ✓ |
| Valine | Branched chain | rs12974412 | 19 | 14151809 | G | A | 0.78 | -0.027 | 0.005 | 4.80E-08 | 115048 | 3 | 30 | 2.59E-04 | ✓ | ✓ | ✓ | ✓ | ✓ | ✓ |
| Valine | Branched chain | rs837616 | 19 | 49365588 | G | A | 0.29 | -0.025 | 0.004 | 1.60E-08 | 115048 | 1 | 32 | 2.78E-04 | ✓ | ✓ | ✓ | ✓ | ✓ | ✓ |
| Valine | Branched chain | rs2238732 | 22 | 18915347 | T | C | 0.04 | -0.062 | 0.010 | 5.80E-10 | 115048 | 1 | 38 | 3.33E-04 | ✓ | NA | ✓ | ✓ | ✓ | ✓ |

Spplementary Table 3: Genetic instruments excluded with associations with more than five metabolites

| Exposure | Subgroup | SNP | Chromosome | Position | Effect allele | Other allele | Effect allele frequency | Beta | Standard error | p-value | Sample size | No. association with NMR |
|---|---|---|---|---|---|---|---|---|---|---|---|---|
| Alanine | Other | rs1047891 | 2 | 211540507 | A | C | 0.32 | -0.028 | 0.004 | 1.60E-10 | 115074 | 12 |
| Alanine | Other | rs1260326 | 2 | 27730940 | C | T | 0.60 | -0.056 | 0.004 | 1.90E-40 | 115074 | 92 |
| Glutamine | Other | rs1260326 | 2 | 27730940 | C | T | 0.60 | 0.079 | 0.004 | 1.60E-81 | 114750 | 92 |
| Glutamine | Other | rs28601761 | 8 | 126500031 | G | C | 0.42 | 0.032 | 0.004 | 1.40E-13 | 114750 | 41 |
| Glutamine | Other | rs117643180 | 17 | 7185779 | A | C | 0.03 | 0.070 | 0.013 | 2.70E-08 | 114750 | 7 |
| Glycine | Other | rs1047891 | 2 | 211540507 | A | C | 0.31 | 0.535 | 0.004 | 1.00E-200 | 114972 | 12 |
| Glycine | Other | rs13107325 | 4 | 103188709 | T | C | 0.07 | -0.049 | 0.007 | 3.00E-13 | 114972 | 33 |
| Glycine | Other | rs28601761 | 8 | 126500031 | G | C | 0.42 | 0.061 | 0.004 | 1.40E-55 | 114972 | 41 |
| Glycine | Other | rs4240624 | 8 | 9184231 | A | G | 0.91 | -0.127 | 0.007 | 2.00E-82 | 114972 | 19 |
| Histidine | Other | rs1047891 | 2 | 211540507 | A | C | 0.31 | -0.070 | 0.004 | 4.00E-55 | 114895 | 12 |
| Histidine | Other | rs11621792 | 14 | 24871926 | T | C | 0.45 | -0.027 | 0.004 | 5.50E-11 | 114895 | 31 |
| Histidine | Other | rs150844304 | 15 | 43726625 | C | A | 0.03 | 0.091 | 0.013 | 3.90E-12 | 114895 | 39 |
| Histidine | Other | rs1883711 | 20 | 39179822 | C | G | 0.03 | 0.087 | 0.012 | 7.70E-13 | 114895 | 67 |
| Tyrosine | Aromatic | rs28601761 | 8 | 126500031 | G | C | 0.42 | -0.038 | 0.004 | 7.30E-19 | 114911 | 41 |
| Tyrosine | Aromatic | rs7979473 | 12 | 121420260 | G | A | 0.61 | -0.062 | 0.004 | 1.80E-47 | 114911 | 19 |
| Tyrosine | Aromatic | rs1800961 | 20 | 43042364 | T | C | 0.03 | -0.073 | 0.012 | 1.30E-09 | 114911 | 24 |
| Tyrosine | Aromatic | rs3747207 | 22 | 44324855 | A | G | 0.22 | 0.029 | 0.005 | 4.80E-09 | 114911 | 6 |
| Isoleucine | Branched chain | rs10184004 | 2 | 165508389 | T | C | 0.41 | -0.030 | 0.004 | 4.00E-13 | 115075 | 8 |
| Isoleucine | Branched chain | rs1260326 | 2 | 27730940 | C | T | 0.60 | -0.047 | 0.004 | 2.30E-29 | 115075 | 92 |
| Isoleucine | Branched chain | rs117643180 | 17 | 7185779 | A | C | 0.03 | -0.101 | 0.013 | 2.90E-15 | 115075 | 7 |
| Leucine | Branched chain | rs1260326 | 2 | 27730940 | C | T | 0.60 | -0.048 | 0.004 | 3.60E-32 | 115074 | 92 |
| Leucine | Branched chain | rs13389219 | 2 | 165528876 | T | C | 0.39 | -0.032 | 0.004 | 2.30E-15 | 115074 | 42 |
| Leucine | Branched chain | rs34894639 | 3 | 135798658 | T | C | 0.23 | -0.026 | 0.005 | 2.50E-08 | 115074 | 9 |
| Leucine | Branched chain | rs117643180 | 17 | 7185779 | A | C | 0.03 | -0.118 | 0.013 | 4.00E-21 | 115074 | 7 |
| Valine | Branched chain | rs1260326 | 2 | 27730940 | C | T | 0.60 | -0.051 | 0.004 | 6.50E-36 | 115048 | 92 |
| Valine | Branched chain | rs1128249 | 2 | 165528624 | T | G | 0.39 | -0.035 | 0.004 | 1.50E-17 | 115048 | 33 |
| Valine | Branched chain | rs117643180 | 17 | 7185779 | A | C | 0.03 | -0.175 | 0.013 | 6.30E-43 | 115048 | 7 |

Spplementary Table 4: Meta-analysed IVW results of genetically predicted amino acids (per SD) on T2D risk

| T2D | | DIAMANTE 2018 (no UKB) | | | | FinnGen (Round 9) | | | | Meta-analysed | | | | |
|---|---|---|---|---|---|---|---|---|---|---|---|---|---|---|
| **Exposure** | **Grouping** | **No. of SNP** | **OR** | **lower CI** | **upper CI** | **No. of SNP** | **OR** | **lower CI** | **upper CI** | **OR** | **lower CI** | **upper CI** | **p-value** | $I^2$ |
| Alanine | Other | 28 | 1.59 | 1.04 | 2.42 | 28 | 1.46 | 1.05 | 2.02 | 1.50 | 1.16 | 1.95 | 0.0020 | 0% |
| Glutamine | Other | 36 | 1.10 | 0.83 | 1.46 | 31 | 1.07 | 0.89 | 1.28 | 1.08 | 0.92 | 1.26 | 0.3531 | 0% |
| Glycine | Other | 35 | 0.90 | 0.76 | 1.06 | 36 | 0.95 | 0.80 | 1.12 | 0.92 | 0.82 | 1.04 | 0.1709 | 0% |
| Histidine | Other | 11 | 1.35 | 0.94 | 1.93 | 11 | 1.29 | 0.96 | 1.75 | 1.32 | 1.04 | 1.66 | 0.0197 | 0% |
| Phenylalanine | Aromatic | 8 | 1.00 | 0.87 | 1.15 | 6 | 1.11 | 1.01 | 1.23 | 1.07 | 0.97 | 1.18 | 0.1895 | 28% |
| Tyrosine | Aromatic | 27 | 0.85 | 0.76 | 0.96 | 25 | 0.96 | 0.81 | 1.14 | 0.89 | 0.80 | 0.99 | 0.0404 | 18% |
| Isoleucine | Branched-chain | 6 | 1.11 | 0.92 | 1.34 | 6 | 1.14 | 0.98 | 1.34 | 1.13 | 1.00 | 1.27 | 0.0478 | 0% |
| Leucine | Branched-chain | 10 | 1.03 | 0.90 | 1.18 | 9 | 1.26 | 1.07 | 1.50 | 1.13 | 0.93 | 1.39 | 0.2205 | 71% |
| Valine | Branched-chain | 15 | 1.18 | 0.81 | 1.71 | 15 | 1.28 | 0.97 | 1.70 | 1.25 | 0.99 | 1.56 | 0.0556 | 0% |

Spplementary Table 4: Effect of genetically predicted amino acids (per SD) on T2D risk (DIAMANTE, Mahajan 2018, no UKB)

| **DIAMANTE (no UKB)** | | | **Inverse variance weighted** | | | | | **Weighted Median** | | | | **MR-Egger** | | | | | **MR-PRESSO** | | | |
|---|---|---|---|---|---|---|---|---|---|---|---|---|---|---|---|---|---|---|---|---|
| **Exposure** | **Grouping** | **No. of SNP** | **OR** | **lower CI** | **upper CI** | **p-value** | **$I^2$** | **OR** | **lower CI** | **upper CI** | **p-value** | **OR** | **lower CI** | **upper CI** | **p-value** | **p-value (Egger's Intercept)** | **OR** | **lower CI** | **upper CI** | **p-value** |
| Alanine | Other | 28 | 1.59 | 1.04 | 2.42 | 0.0335 | 95% | 1.07 | 0.91 | 1.26 | 0.4115 | 0.74 | 0.26 | 2.11 | 0.5833 | 0.13 | 1.19 | 0.97 | 1.45 | 0.1162 |
| Glutamine | Other | 36 | 1.10 | 0.83 | 1.46 | 0.5107 | 96% | 1.03 | 0.94 | 1.12 | 0.5381 | 1.14 | 0.75 | 1.75 | 0.5461 | 0.82 | 1.04 | 0.95 | 1.13 | 0.4253 |
| Glycine | Other | 35 | 0.90 | 0.76 | 1.06 | 0.2010 | 80% | 0.93 | 0.82 | 1.04 | 0.2056 | 1.17 | 0.85 | 1.61 | 0.3411 | 0.06 | 0.94 | 0.87 | 1.03 | 0.2007 |
| Histidine | Other | 11 | 1.35 | 0.94 | 1.93 | 0.1002 | 84% | 1.08 | 0.88 | 1.32 | 0.4507 | 0.48 | 0.23 | 0.99 | 0.0460 | 0.01 | 1.09 | 0.93 | 1.28 | 0.2938 |
| Phenylalanine | Aromatic | 8 | 1.00 | 0.87 | 1.15 | 0.9649 | 28% | 1.06 | 0.93 | 1.21 | 0.3892 | 1.08 | 0.82 | 1.41 | 0.6123 | 0.57 | NA | NA | NA | NA |
| Tyrosine | Aromatic | 27 | 0.85 | 0.76 | 0.96 | 0.0099 | 61% | 0.87 | 0.78 | 0.97 | 0.0103 | 0.89 | 0.74 | 1.07 | 0.2176 | 0.57 | 0.91 | 0.84 | 0.98 | 0.0226 |
| Isoleucine | Branched-chain | 6 | 1.11 | 0.92 | 1.34 | 0.2737 | 25% | 1.07 | 0.88 | 1.31 | 0.4887 | 1.92 | 1.14 | 3.21 | 0.0135 | 0.09 | NA | NA | NA | NA |
| Leucine | Branched-chain | 10 | 1.03 | 0.90 | 1.18 | 0.6671 | 0% | 1.04 | 0.88 | 1.23 | 0.6261 | 1.16 | 0.83 | 1.62 | 0.4085 | 0.47 | NA | NA | NA | NA |
| Valine | Branched-chain | 15 | 1.18 | 0.81 | 1.71 | 0.3860 | 92% | 1.04 | 0.91 | 1.20 | 0.5477 | 0.74 | 0.39 | 1.40 | 0.3713 | 0.11 | 1.03 | 0.92 | 1.14 | 0.6560 |

Spplementary Table 5: Effect of genetically predicted amino acids (per SD) on T2D risk (FinnGen, Round 9)

| FinnGen (Round 9) | | | Inverse variance weighted | | | | | Weighted median | | | | MR-Egger | | | | | MR-PRESSO | | | |
|---|---|---|---|---|---|---|---|---|---|---|---|---|---|---|---|---|---|---|---|---|
| **Exposure** | **Grouping** | **No. of SNP** | **OR** | **lower CI** | **upper CI** | **P value** | **$I^2$** | **OR** | **lower CI** | **upper CI** | **P value** | **OR** | **lower CI** | **upper CI** | **P value** | **P value (Egger's Intercept)** | **OR** | **lower CI** | **upper CI** | **P value** |
| Alanine | Other | 28 | 1.46 | 1.05 | 2.02 | 0.0230 | 95% | 1.09 | 0.95 | 1.25 | 0.2033 | 0.97 | 0.40 | 2.35 | 0.9381 | 0.34 | 1.14 | 1.00 | 1.31 | 0.0662 |
| Glutamine | Other | 31 | 1.07 | 0.89 | 1.28 | 0.4959 | 94% | 0.95 | 0.89 | 1.02 | 0.1311 | 0.99 | 0.75 | 1.30 | 0.9375 | 0.46 | 1.07 | 0.99 | 1.17 | 0.1120 |
| Glycine | Other | 36 | 0.95 | 0.80 | 1.12 | 0.5059 | 87% | 0.98 | 0.90 | 1.07 | 0.6570 | 1.31 | 0.98 | 1.75 | 0.0818 | 0.02 | 0.98 | 0.91 | 1.07 | 0.7013 |
| Histidine | Other | 11 | 1.29 | 0.96 | 1.75 | 0.0962 | 85% | 1.15 | 0.95 | 1.39 | 0.1435 | 0.47 | 0.27 | 0.81 | 0.0236 | 3.64E-03 | 1.23 | 1.01 | 1.50 | 0.0790 |
| Phenylalanine | Aromatic | 6 | 1.11 | 1.01 | 1.23 | 0.0392 | 0% | 1.09 | 0.97 | 1.22 | 0.1389 | 0.99 | 0.82 | 1.20 | 0.9270 | 0.24 | NA | NA | NA | NA |
| Tyrosine | Aromatic | 25 | 0.96 | 0.81 | 1.14 | 0.6486 | 84% | 0.96 | 0.86 | 1.06 | 0.4252 | 1.06 | 0.78 | 1.44 | 0.7052 | 0.44 | 0.97 | 0.88 | 1.06 | 0.4652 |
| Isoleucine | Branched-chain | 6 | 1.14 | 0.98 | 1.34 | 0.0958 | 0% | 1.16 | 0.96 | 1.41 | 0.1283 | 1.45 | 0.91 | 2.30 | 0.1947 | 0.35 | NA | NA | NA | NA |
| Leucine | Branched-chain | 9 | 1.26 | 1.07 | 1.50 | 0.0070 | 22% | 1.34 | 1.09 | 1.66 | 0.0057 | 1.66 | 1.00 | 2.76 | 0.0905 | 0.30 | NA | NA | NA | NA |
| Valine | Branched-chain | 15 | 1.28 | 0.97 | 1.70 | 0.0811 | 87% | 1.18 | 1.02 | 1.37 | 0.0292 | 0.71 | 0.41 | 1.22 | 0.2388 | 0.03 | 1.26 | 1.09 | 1.45 | 0.0082 |

Spplementary Table 6: Effect of genetically predicted amino acids (per SD) on fasting insulin (FI, log transformed)

| FI (MAGIC) | | | Inverse variance weighted | | | | Weighted Median | | | MR-Egger | | | | MR-PRESSO | | |
|---|---|---|---|---|---|---|---|---|---|---|---|---|---|---|---|---|
| **Exposure** | **Grouping** | **No. of SNP** | **Beta** | **SE** | **P value** | **$I^2$** | **Beta** | **SE** | **P value** | **Beta** | **SE** | **P value** | **P value (Egger's Intercept)** | **Beta** | **SE** | **P value** |
| Alanine | Other | 30 | -0.059 | 0.017 | 0.0004 | 52% | -0.060 | 0.018 | 0.0010 | -0.035 | 0.044 | 0.4361 | 0.55 | -0.051 | 0.014 | 0.0011 |
| Glutamine | Other | 41 | 0.002 | 0.016 | 0.9050 | 82% | 0.020 | 0.010 | 0.0367 | 0.028 | 0.023 | 0.2385 | 0.13 | -0.005 | 0.017 | 0.7525 |
| Glycine | Other | 43 | -0.027 | 0.025 | 0.2734 | 86% | -0.011 | 0.015 | 0.4652 | 0.034 | 0.050 | 0.5039 | 0.17 | -0.016 | 0.012 | 0.1795 |
| Histidine | Other | 13 | 0.044 | 0.066 | 0.5039 | 93% | -0.001 | 0.030 | 0.9787 | -0.109 | 0.175 | 0.5469 | 0.37 | 0.034 | 0.030 | 0.2938 |
| Phenylalanine | Aromatic | 8 | -0.032 | 0.018 | 0.0838 | 44% | -0.040 | 0.017 | 0.0166 | -0.056 | 0.039 | 0.1997 | 0.50 | NA | NA | NA |
| Tyrosine | Aromatic | 30 | -0.004 | 0.016 | 0.7822 | 62% | -0.005 | 0.014 | 0.7273 | -0.014 | 0.025 | 0.5614 | 0.60 | -0.012 | 0.011 | 0.3082 |
| Isoleucine | Branched-chain | 7 | -0.069 | 0.019 | 0.0004 | 0% | -0.061 | 0.024 | 0.0124 | -0.080 | 0.055 | 0.2071 | 0.85 | NA | NA | NA |
| Leucine | Branched-chain | 11 | -0.051 | 0.017 | 0.0021 | 0% | -0.047 | 0.020 | 0.0215 | -0.038 | 0.039 | 0.3484 | 0.73 | NA | NA | NA |
| Valine | Branched-chain | 17 | 0.008 | 0.044 | 0.8613 | 92% | -0.032 | 0.017 | 0.0634 | -0.136 | 0.080 | 0.1083 | 0.05 | -0.007 | 0.023 | 0.7579 |

Spplementary Table 7: Effect of genetically predicted amino acids (per SD) on glycated hemoglobin (HbA1c, %)

| HbA1c (MAGIC) | | | Inverse variance weighted | | | | Weighted Median | | | MR-Egger | | | | MR-PRESSO | | |
|---|---|---|---|---|---|---|---|---|---|---|---|---|---|---|---|---|
| Exposure | Grouping | No. of SNP | Beta | SE | P value | $I^2$ | Beta | SE | P value | Beta | SE | P value | P value (Egger's Intercept) | Beta | SE | P value |
| Alanine | Other | 30 | 0.065 | 0.023 | 0.0053 | 89% | 0.019 | 0.014 | 0.1951 | -0.016 | 0.060 | 0.7909 | 0.16 | 0.042 | 0.011 | 0.0009 |
| Glutamine | Other | 41 | 0.013 | 0.011 | 0.2509 | 83% | 0.004 | 0.006 | 0.5276 | 0.015 | 0.017 | 0.3868 | 0.89 | 0.004 | 0.007 | 0.5754 |
| Glycine | Other | 43 | -0.004 | 0.010 | 0.6899 | 60% | -0.006 | 0.009 | 0.5074 | 0.019 | 0.019 | 0.3385 | 0.19 | -0.008 | 0.008 | 0.3146 |
| Histidine | Other | 13 | 0.028 | 0.020 | 0.1597 | 66% | 0.010 | 0.019 | 0.5905 | -0.061 | 0.047 | 0.2174 | 0.06 | 0.008 | 0.014 | 0.6037 |
| Phenylalanine | Aromatic | 8 | 0.023 | 0.012 | 0.0647 | 48% | 0.021 | 0.011 | 0.0539 | 0.011 | 0.027 | 0.7061 | 0.63 | NA | NA | NA |
| Tyrosine | Aromatic | 30 | -0.014 | 0.012 | 0.2249 | 71% | -0.013 | 0.009 | 0.1540 | -0.027 | 0.018 | 0.1459 | 0.36 | -0.010 | 0.009 | 0.251 |
| Isoleucine | Branched-chain | 7 | -0.008 | 0.014 | 0.5716 | 24% | -0.018 | 0.016 | 0.2665 | -0.054 | 0.039 | 0.2246 | 0.26 | NA | NA | NA |
| Leucine | Branched-chain | 11 | -0.003 | 0.012 | 0.8063 | 23% | -0.010 | 0.013 | 0.4465 | -0.048 | 0.025 | 0.0890 | 0.08 | NA | NA | NA |
| Valine | Branched-chain | 17 | -0.002 | 0.013 | 0.8813 | 59% | -0.011 | 0.011 | 0.3249 | -0.036 | 0.025 | 0.1665 | 0.13 | -0.007 | 0.010 | 0.4962 |

Spplementary Table 8: Effect of genetically predicted amino acids (per SD) on fasting glucose (FG, mmol/L)

| FG (MAGIC) | | | Inverse variance weighted | | | | Weighted Median | | | MR-Egger | | | | MR-PRESSO | | |
|---|---|---|---|---|---|---|---|---|---|---|---|---|---|---|---|---|
| **Exposure** | **Grouping** | **No. of SNP** | **Beta** | **SE** | **P value** | **$I^2$** | **Beta** | **SE** | **P value** | **Beta** | **SE** | **P value** | **P value (Egger's Intercept)** | **Beta** | **SE** | **P value** |
| Alanine | Other | 30 | 0.142 | 0.043 | 0.0010 | 95% | 0.071 | 0.019 | 0.0002 | 0.149 | 0.115 | 0.2060 | 0.95 | 0.079 | 0.017 | 0.0002 |
| Glutamine | Other | 41 | 0.004 | 0.021 | 0.8299 | 91% | -0.041 | 0.010 | 1.40E-05 | -0.024 | 0.030 | 0.4210 | 0.19 | 0.018 | 0.014 | 0.2351 |
| Glycine | Other | 43 | 0.022 | 0.018 | 0.2267 | 80% | 0.006 | 0.012 | 0.6246 | 0.012 | 0.037 | 0.7535 | 0.76 | 0.007 | 0.012 | 0.5632 |
| Histidine | Other | 13 | 0.124 | 0.077 | 0.1085 | 96% | -0.018 | 0.028 | 0.5191 | -0.161 | 0.193 | 0.4232 | 0.14 | 0.098 | 0.035 | 0.0276 |
| Phenylalanine | Aromatic | 8 | -0.017 | 0.024 | 0.4870 | 75% | -0.017 | 0.015 | 0.2440 | -0.025 | 0.054 | 0.6620 | 0.87 | -0.025 | 0.018 | 0.2177 |
| Tyrosine | Aromatic | 30 | -0.009 | 0.023 | 0.6798 | 86% | -0.011 | 0.013 | 0.3949 | 0.009 | 0.036 | 0.8121 | 0.52 | -0.003 | 0.010 | 0.7758 |
| Isoleucine | Branched-chain | 7 | 0.034 | 0.040 | 0.3973 | 82% | 0.023 | 0.024 | 0.3335 | -0.044 | 0.120 | 0.7280 | 0.52 | 0.029 | 0.014 | 0.1125 |
| Leucine | Branched-chain | 11 | 0.037 | 0.024 | 0.1258 | 63% | 0.025 | 0.018 | 0.1727 | 0.008 | 0.058 | 0.8983 | 0.60 | 0.018 | 0.010 | 0.1102 |
| Valine | Branched-chain | 17 | 0.038 | 0.021 | 0.0664 | 70% | 0.023 | 0.015 | 0.1282 | -0.033 | 0.036 | 0.3737 | 0.04 | 0.019 | 0.012 | 0.1453 |

Spplementary Table 9: Effect of genetically predicted amino acids (per SD) on 2-hour glucose (2hGlu, mmol/L)

| 2hGlu (MAGIC) | | | Inverse variance weighted | | | | Weighted Median | | | MR-Egger | | | | MR-PRESSO | | |
|---|---|---|---|---|---|---|---|---|---|---|---|---|---|---|---|---|
| **Exposure** | **Grouping** | **No. of SNP** | **Beta** | **SE** | **P value** | **$I^2$** | **Beta** | **SE** | **P value** | **Beta** | **SE** | **P value** | **P value (Egger's Intercept)** | **Beta** | **SE** | **P value** |
| Alanine | Other | 30 | 0.170 | 0.095 | 0.0730 | 76% | 0.126 | 0.072 | 0.0813 | -0.009 | 0.240 | 0.9687 | 0.42 | 0.076 | 0.064 | 0.2441 |
| Glutamine | Other | 41 | -0.006 | 0.046 | 0.8890 | 64% | -0.065 | 0.037 | 0.0759 | -0.030 | 0.067 | 0.6581 | 0.63 | -0.026 | 0.030 | 0.3907 |
| Glycine | Other | 43 | -0.102 | 0.048 | 0.0343 | 36% | -0.080 | 0.060 | 0.1852 | 2.03E-04 | 0.097 | 0.9983 | 0.23 | -0.083 | 0.043 | 0.0612 |
| Histidine | Other | 13 | -0.135 | 0.127 | 0.2874 | 66% | 0.014 | 0.111 | 0.8985 | 0.161 | 0.335 | 0.6393 | 0.36 | -0.003 | 0.067 | 0.9617 |
| Phenylalanine | Aromatic | 8 | -0.032 | 0.089 | 0.7179 | 57% | -0.048 | 0.073 | 0.5090 | -0.197 | 0.182 | 0.3194 | 0.34 | NA | NA | NA |
| Tyrosine | Aromatic | 30 | 0.002 | 0.054 | 0.9679 | 47% | 0.020 | 0.060 | 0.7408 | 0.017 | 0.085 | 0.8472 | 0.83 | -0.014 | 0.049 | 0.7798 |
| Isoleucine | Branched-chain | 7 | 0.043 | 0.086 | 0.6184 | 11% | -0.072 | 0.102 | 0.4805 | -0.340 | 0.229 | 0.1973 | 0.13 | NA | NA | NA |
| Leucine | Branched-chain | 11 | 0.026 | 0.078 | 0.7430 | 22% | -0.032 | 0.087 | 0.7121 | -0.293 | 0.163 | 0.1050 | 0.06 | NA | NA | NA |
| Valine | Branched-chain | 17 | 0.006 | 0.083 | 0.9399 | 58% | -0.051 | 0.071 | 0.4785 | -0.143 | 0.163 | 0.3946 | 0.31 | -0.011 | 0.043 | 0.8020 |
